# Biallelic variation in the choline and ethanolamine transporter *FLVCR1* underlies a pleiotropic disease spectrum from adult neurodegeneration to severe developmental disorders

**DOI:** 10.1101/2024.02.09.24302464

**Authors:** Daniel G. Calame, Jovi Huixin Wong, Puravi Panda, Dat Tuan Nguyen, Nancy C.P. Leong, Riccardo Sangermano, Sohil G. Patankar, Mohamed Abdel-Hamid, Lama AlAbdi, Sylvia Safwat, Kyle P. Flannery, Zain Dardas, Jawid M. Fatih, Chaya Murali, Varun Kannan, Timothy E. Lotze, Isabella Herman, Farah Ammouri, Brianna Rezich, Stephanie Efthymiou, Shahryar Alavi, David Murphy, Zahra Firoozfar, Mahya Ebrahimi Nasab, Amir Bahreini, Majid Ghasemi, Nourelhoda A. Haridy, Hamid Reza Goldouzi, Fatemeh Eghbal, Ehsan Ghayoor Karimiani, Varunvenkat M. Srinivasan, Vykuntaraju K. Gowda, Haowei Du, Shalini N. Jhangiani, Zeynep Coban-Akdemir, Dana Marafi, Lance Rodan, Sedat Isikay, Jill A. Rosenfeld, Subhadra Ramanathan, Michael Staton, Kerby C. Oberg, Robin D. Clark, Catharina Wenman, Sam Loughlin, Ramy Saad, Tazeen Ashraf, Alison Male, Shereen Tadros, Reza Boostani, Ghada M.H. Abdel-Salam, Maha Zaki, Ebtesam Abdalla, M. Chiara Manzini, Davut Pehlivan, Jennifer E. Posey, Richard A. Gibbs, Henry Houlden, Fowzan S. Alkuraya, Kinga Bujakowska, Reza Maroofian, James R. Lupski, Long Nam Nguyen

## Abstract

*FLVCR1* encodes Feline leukemia virus subgroup C receptor 1 (FLVCR1), a solute carrier (SLC) transporter within the Major Facilitator Superfamily. FLVCR1 is a widely expressed transmembrane protein with plasma membrane and mitochondrial isoforms implicated in heme, choline, and ethanolamine transport. While *Flvcr1* knockout mice die *in utero* with skeletal malformations and defective erythropoiesis reminiscent of Diamond-Blackfan anemia, rare biallelic pathogenic *FLVCR1* variants are linked to childhood or adult-onset neurodegeneration of the retina, spinal cord, and peripheral nervous system.

We ascertained from research and clinical exome sequencing 27 individuals from 20 unrelated families with biallelic ultra-rare missense and predicted loss-of-function (pLoF) *FLVCR1* variant alleles. We characterize an expansive *FLVCR1* phenotypic spectrum ranging from adult-onset retinitis pigmentosa to severe developmental disorders with microcephaly, reduced brain volume, epilepsy, spasticity, and premature death. The most severely affected individuals, including three individuals with homozygous pLoF variants, share traits with *Flvcr1* knockout mice and Diamond-Blackfan anemia including macrocytic anemia and congenital skeletal malformations. Pathogenic *FLVCR1* missense variants primarily lie within transmembrane domains and reduce choline and ethanolamine transport activity compared with wild-type *FLVCR1* with minimal impact on FLVCR1 stability or subcellular localization. Several variants disrupt splicing in a mini-gene assay which may contribute to genotype-phenotype correlations. Taken together, these data support an allele-specific gene dosage model in which phenotypic severity reflects residual FLVCR1 activity. This study expands our understanding of Mendelian disorders of choline and ethanolamine transport and demonstrates the importance of choline and ethanolamine in neurodevelopment and neuronal homeostasis.

## Text

Solute transport across lipid bilayers is critical for biological homeostasis and requires specialized transmembrane proteins^1^. The largest group of membrane transport proteins is the solute carrier (SLC) superfamily. The superfamily consists of 458 genes divided into 65 families^1^. SLC proteins transport a vast assortment of solutes including amino acids, sugars, ions, nucleotides, vitamins, and neurotransmitters. Genetic variation in SLC genes influences common diseases, metabolic traits, and an expanding spectrum of Mendelian disorders^2–4^. As solute transport disorders are responsive to solute supplementation, dietary interventions, and gene therapies in preclinical models and humans, they are attractive therapeutic targets^5–7^. Yet, the function of most SLC genes in human biology remains poorly characterized.

One SLC family member associated with Mendelian neurodegenerative disorders is *FLVCR1* (also known as *SLC49A1* and *MFSD7B*). First recognized as the receptor for Feline Leukemia Virus Subgroup C (FeLV-C), a retrovirus which causes aplastic anemia in cats^8^, *FLVCR1* is highly conserved and encodes two isoforms in humans: a full-length 555 amino acid (aa) plasma membrane isoform FLVCR1a and a small mitochondrial membrane isoform FLVCR1b (aa 277-555)^9^. FLVCR1 was initially described as a heme exporter in erythroid cells and other cell lineages^10^. An essential requirement for FLVCR1 was subsequently demonstrated through knockout of the mouse ortholog *Flvcr1*^11^. Germline knockout of *Flvcr1* results in embryonic lethality, absent erythropoiesis, craniofacial and limb malformations, and hepatic iron accumulation, whereas neonatal deletion causes severe macrocytic anemia^11^. Both FLVCR1 isoforms are required for murine viability; mice retaining FLVCR1b but lacking FLVCR1a have normal erythropoiesis but develop hemorrhages, edema, and skeletal malformations^9^. The physiologic significance of FLVCR1 heme transport has been called in question especially considering recent evidence that FLVCR1 is a choline and ethanolamine transporter^4,12–16^.

The severe anemia and craniofacial, limb and digital malformations in *Flvcr1* knockout mice resemble humans with Diamond-Blackfan anemia (DBA) [MIM: 105650]^11,17^. A further link between *FLVCR1* and DBA was suggested by the observation that *FLVCR1* splicing is dysregulated in erythroid cells from patients with DBA^18^. However, biallelic variation in *FLVCR1* has not been identified in individuals with DBA, and DBA is now recognized as primarily a disorder of ribosome biogenesis^17^. Instead, biallelic pathogenic variation in *FLVCR1* causes rare recessive neurodegenerative disorders of the retina, spinal cord, and peripheral nerves: posterior column ataxia with retinitis pigmentosa (PCARP) [MIM: 609033], isolated retinitis pigmentosa (RP), and hereditary sensory and autonomic neuropathy (HSAN)^19–21^. *FLVCR1*-related neurodegenerative disorders can manifest from childhood into adulthood. To date, at least 39 individuals from 22 families have been identified with FLVCR1-related neurodegeneration; most have biallelic *FLVCR1* missense variants, and predicted loss-of-function variants have only been identified *in trans* with missense variants (**Supplemental Table S1**). Despite the severe multi-organ consequences of *Flvcr1* knockout in mice, extra-neurological findings or severe developmental disorders have only been described in one individual with biallelic *FLVCR1* variants, a compound heterozygote (c.574T>C p.Cys192Arg; c.610del p.Met204Cysfs*56) with severe developmental disabilities, chronic macrocytic anemia, liver disease, self-mutilation, and sensory neuropathy^21^. The precise reason(s) for the apparent discrepancies between human and mouse *FLVCR1*-related phenotypes remains unclear but could suggest that *FLVCR1* knockout is incompatible with human life.

The index case who initiated this study is a male child with severe developmental delay, epileptic encephalopathy, and microcephaly (Individual 1, **Fig. 1**, **Fig. 2A-E**, **Table 1**). He was born to non-consanguineous parents from a country in South Asia. He had a history of infantile spasms, self-mutilation, osteomyelitis, and absent sensory nerve responses on nerve conduction studies. Brain magnetic resonance imaging (MRI) showed corpus callosum thinning, brainstem and pontine thinning, prominent extra-axial fluid spaces, and reduced white matter volume (**Fig. 2A-E**). Uric acid and purine levels, measured to assess for Lesch-Nyhan syndrome [MIM: 300322] due to the history of severe self-injury, were normal. Clinical trio exome sequencing (cES) did not identify any variants in *HPRT1* nor pathogenic variants in known disease genes. Hence, these cES data underwent research reanalysis through the Baylor College of Medicine Genomics Research Elucidates the Genetics of Rare diseases (BCM-GREGoR) which prioritized the homozygous *FLVCR1* missense variant c.1390G>A p.G464S given the overlap between the patient’s known sensory neuropathy and known *FLVCR1*-related disorders.

**Figure 1:** Pedigrees of families with *FLVCR1*-related developmental and neurodegenerative disorders. **Per medrxiv requirements, this figure was removed but is available upon request to the corresponding authors.** Pedigrees of families with *FLVCR1*-related diseases. Genotype is indicated above each pedigree and below each family member. *A* indicates wild-type *FLVCR1* allele, *a* indicates *FLVCR1* variant allele 1, and *a’* indicates *FLVCR1* variant allele 2; specific variants differ from one pedigree to the next. Red shading indicates severe *FLVCR1*-related disease (microcephaly, brain atrophy, severe developmental disabilities), whereas light blue shading indicates relatively milder neurological *FLVCR1*-related disease (posterior column ataxia with retinitis pigmentosa, hereditary sensory and autonomic neuropathy, isolated retinitis pigmentosa, or hereditary spastic paraplegia). Gray shading indicates individuals suspected of possibly having a different genetic syndrome characterized by neonatal demise, hypotonia, and arthrogryposis.

**Figure 2:**
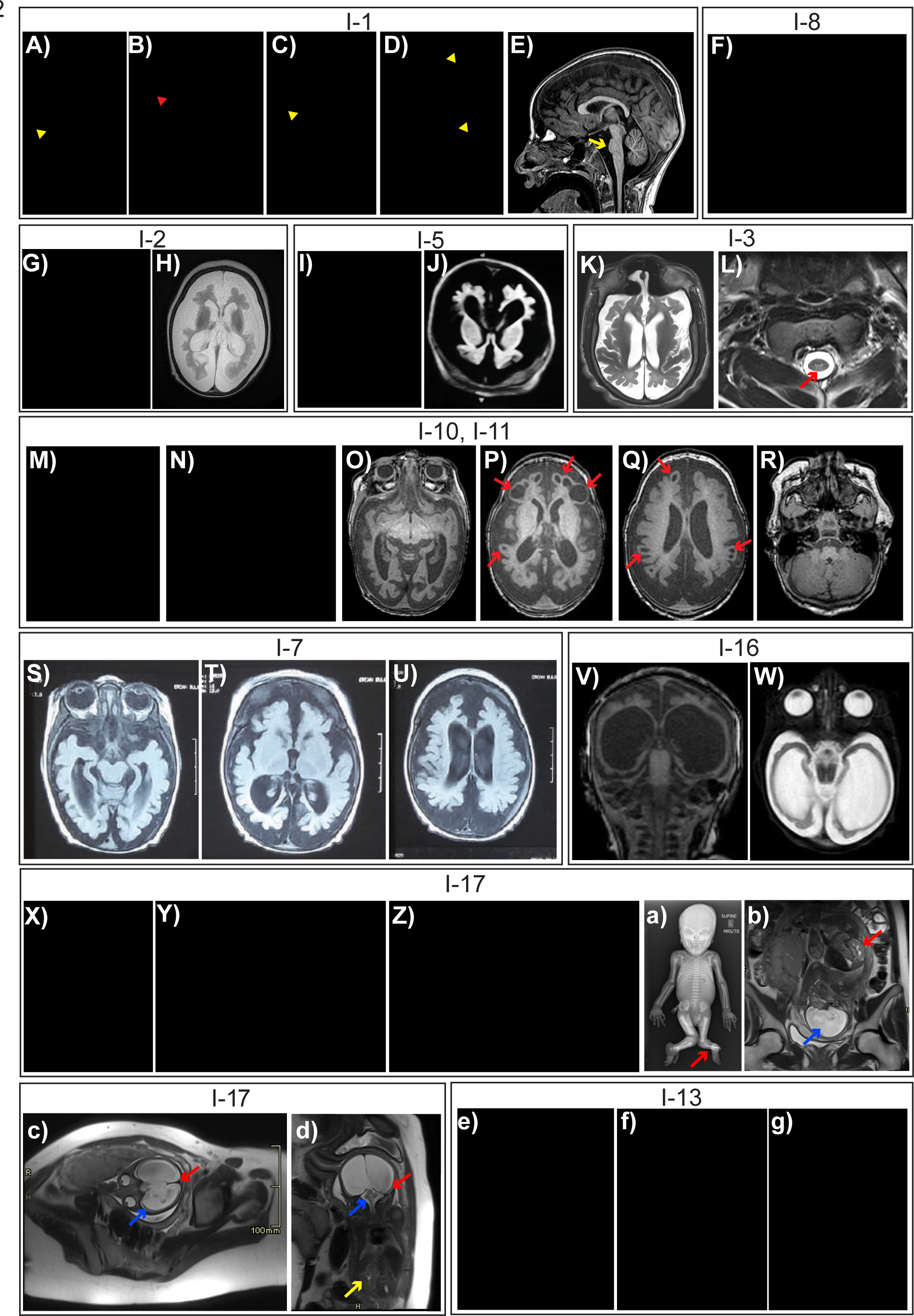
Phenotypic features of *FLVCR1*-related developmental and neurodegenerative disorders. Photographs and magnetic resonance imaging (MRI) from each family are divided into separate boxes bordered by thin black lines. **Per medrxiv requirements, patient photographs have been removed but are available upon request to the corresponding authors.** A-E) Images of Individual 1 (I-1), Family 1 with homozygous *FLVCR1* p.G464S variant. A-D show examples of self-mutilation including foot, ear, and arm ulcerations (yellow arrowheads) and digital amputation (red arrowhead). E shows sagittal T1-weighted brain MRI. Pontine thinning is indicated with yellow arrow. **F)** Image of Individual 8 (I-8), Family 7 with homozygous *FLVCR1* p.G464S variant. Self-mutilation and digital amputation of the hand is shown. **G, H)** Images of Individual 2 (I-2), Family 2. G shows patient with triangular facies, microcephaly and severe developmental delay. H shows axial T2-weighted brain MRI demonstrating severe reduction in cerebral brain volume and a simplified gyral pattern. **I, J)** Images of Individual 5 (I-5), Family 5. I shows patient with microcephaly, broad nasal bridge, widely spaced eyes, and severe developmental delay. J shows axial T1-weighted brain MRI, demonstrating severe reduction in cerebral brain volume and a simplified gyral pattern. **K, L)** Images of Individual 3 (I-3), Family 3. K shows axial T2-weighted brain MRI with severe reduction in cerebral brain volume. L shows T2-weighted cervical spine MRI demonstrating posterior column T2 signal hyperintensity (red arrow). **M-R**) Images of Individuals 10 and 11 (I-10 and I-11), Family 8. M and N show individuals 10 and 11. Photographs demonstrating microcephaly, severe developmental delay, broad nasal bridge, and tracheostomy secondary to respiratory failure. O-R show axial T1-weighted brain MRI from Individual 11 demonstrating reduction in cerebral brain volume with relative cerebellar sparing and cystic encephalomalacia (red arrows). **S-U)** Images of Individual 7 (I-7), Family 6. Images show axial T2-weighted brain MRI with reduction in cerebral brain volume. **V, W**) Images of Individual 16 (I-16), Family 11. V and W show coronal T2-weighted brain MRI and axial T1-weighted brain MRI respectively demonstrating a severe reduction in brain volume with hydrocephalus *ex vacuo*. X-d) Images of Individual 17 (I-17), Family 12. X shows irregular undulation of the ear lobe. Y shows the left foot with ankle dislocation and absence of the great toe, 2^nd^ toe, and 3^rd^ toe. Z shows the right foot demonstrating cleft foot, absence of the 2^nd^ toe, syndactyly of toes 3-4, and proximal displacement of the great toe. a is a full body radiograph demonstrating ten ribs, long limbs, absent left tibia (red arrow), and bilateral ectrodactyly of the feet. b-d are fetal T2-weighted MRI images performed at 28 weeks gestation. b shows hydrocephalus with absence of midline structures (blue arrow) and hypertrophic kidney with pelviectasis (red arrow). c shows hydrocephalus with absence of some midline structures (red arrow) and thinning of the cortical mantle (blue arrow). d shows severe hydrocephalus with thinning of the cortical mantle (red arrow), small cerebellum and brainstem (blue arrow), and left kidney hypertrophy with absence of the right kidney (yellow arrow). e-g) Images of Individual 13 (I-13), Family 9. Postmortem images of Individual 13 (c-e) show joint contractures, long tapering figures. absent right thumb, hypoplastic left thumb, microcephaly, and cleft lip and palate.

**Table 1:**
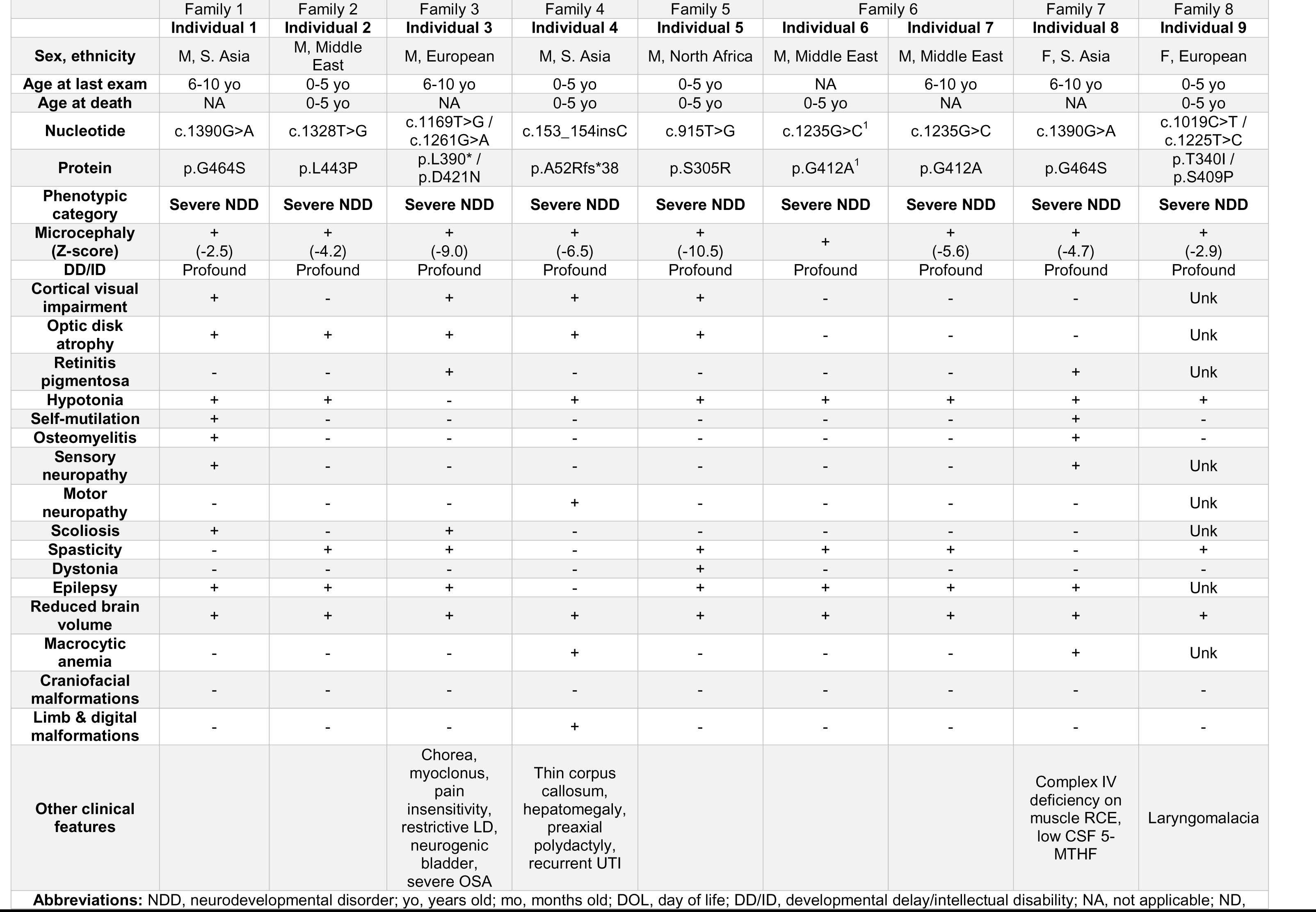
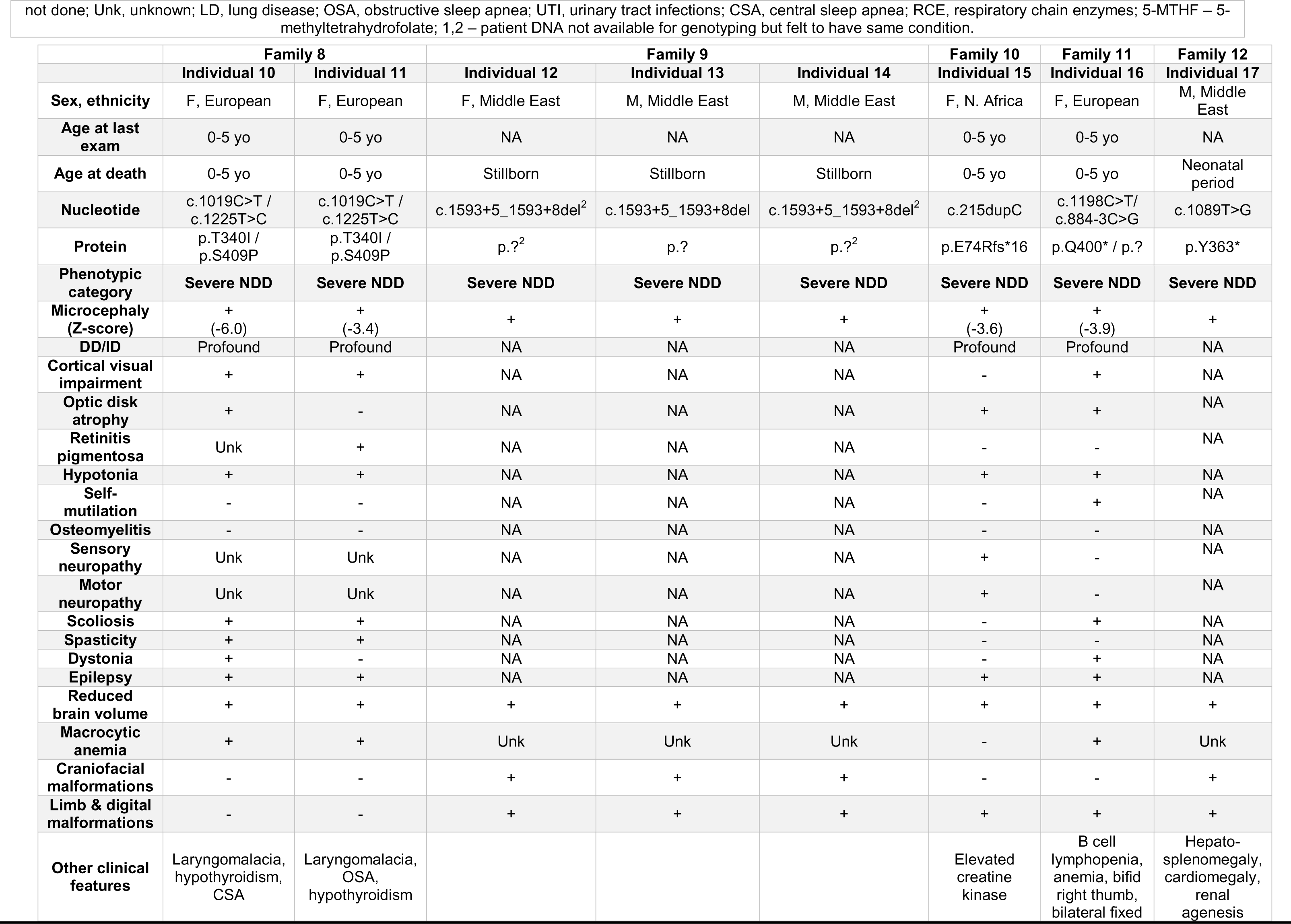
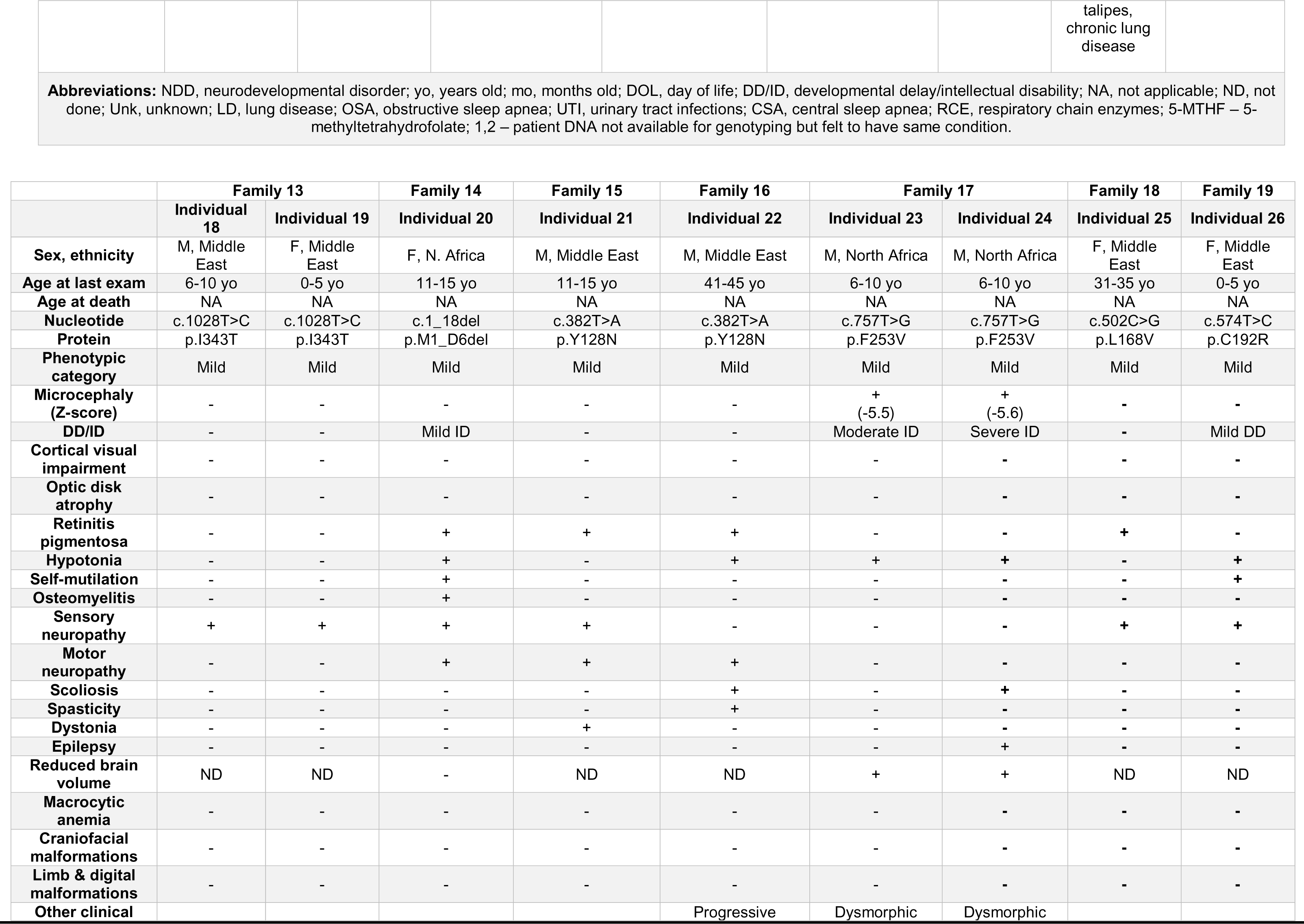
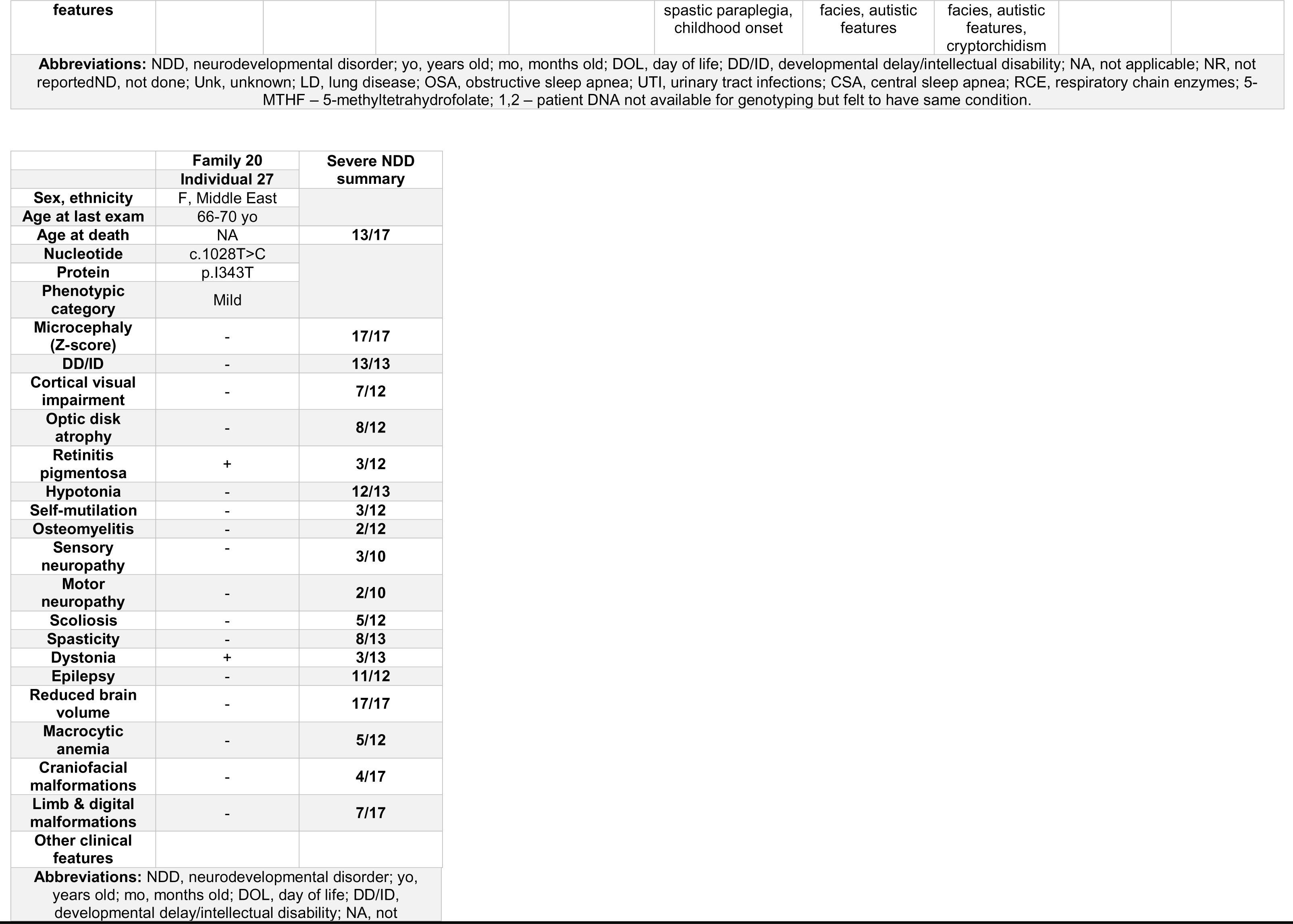
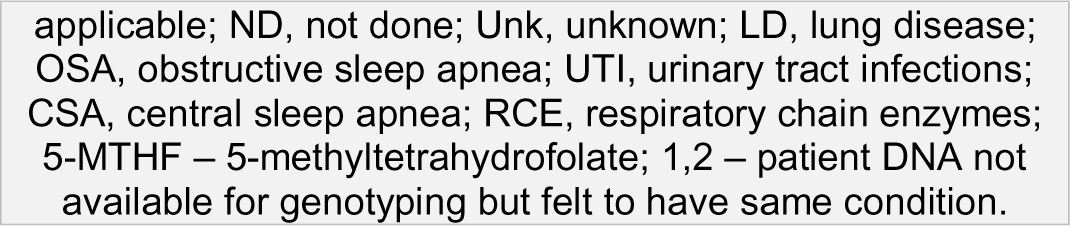
Phenotypic summary of individuals with *FLVCR1*-related developmental and neurodegenerative disorders

To comprehensively characterize the phenotypic spectrum of *FLVCR1*-related disorders in humans, we reanalyzed ES and genome sequencing (GS) data from the 29,766 individuals within the BCM-GREGoR and Baylor Genetics clinical diagnostic databases^22^, utilized the online matchmaking program GeneMatcher^23,24^, and searched other research and diagnostic lab datasets. We identified 27 individuals from 20 unrelated families with neurological disorders and biallelic *FLVCR1* variants (**Fig. 1**, **Table 1**). A pedigree was not available for Individual 27, an elderly woman with isolated retinitis pigmentosa and dystonia. Unlike prior reports describing biallelic *FLVCR1* variants in children or adults with neurodegenerative disorders, Individuals 1-17 had severe developmental disorders. Notably, three individuals with severe developmental disorders had homozygous *FLVCR1* pLoF variants (Individuals 4, 15, and 17). Individuals 18-27 had childhood or adult-onset neurodegenerative disorders including PCARP, hereditary spastic paraplegia, and RP (**Table 1**). Most families (14/20) are consanguineous by clinical history. In Families 1 and 7, the same homozygous missense variant c.1390G>A p.G464S was identified. Both families originate in the same country in South Asia and are unrelated. Neither family are consanguineous by clinical history. Absence-of-heterozygosity (AOH), a surrogate measure from ES data for runs-of-homozygosity (ROH), was calculated for Family 1; total autosomal AOH was 150.1 Mb, and the variant was surrounded by a 4.9 Mb AOH block (**Supplemental Fig. 1**)^25^. These data suggest *FLVCR1*: c.1390G>A p.G464S may represent a South Asian founder allele. Compound heterozygous variants were identified in the three non-consanguineous families and segregated according to Mendelian expectations within Family 8. All clinical and research data were acquired in accordance with ethical standards upon informed consent and with the approval of the collaborative institutional review boards (**Supplemental Methods**).

The phenotypic features of the 27 individuals are summarized in **Table 1**, **Fig. 2**, **and Supplemental Table 1**. Individuals 1-17 exhibited severe developmental delay: all were non-verbal and achieved no developmental motor milestones. All were microcephalic (median Z-score -4.45, range -2.5 to -10.5) and had reduced brain volume on brain magnetic resonance imaging (MRI). Individual 16 was microcephalic at birth (29 cm, Z-score -3.9) but was normocephalic at 3 years old (50.2 cm, Z-score +1.05). Brain MRI findings varied considerably. Some individuals had only a mild reduction in white matter volume, corpus callosum thinning, and/or pontine and brainstem thinning (**Fig. 2E**). Others had a severe reduction in brain volume with simplified gyral pattern (**Fig. 2H, J, K**). In Family 8, cystic encephalomalacia was seen in all three affected siblings (**Fig. 2O-R**); cystic encephalomalacia was detected on fetal MRI suggesting a very early developmental defect. Individuals 16 and 17 had only a thin rim of cerebral cortex reminiscent of hydrancephaly. Another recurring MRI finding observed in Individuals 1 and 3 was T2 hyperintensity of the posterior columns on spine MRI as previously reported in PCARP (**Fig. 2L**). Premature death before adulthood was common (14/17). Hypotonia and epilepsy were nearly universal in individuals who survived the neonatal period. Other common traits include cortical visual impairment, optic disk atrophy, and spasticity. Features previously associated with *FLVCR1* including RP and sensory neuropathy were observed but uncommon; this could represent age-dependent penetrance due to the young age of the cohort or under-ascertainment. Individuals 1 and 8 with the homozygous p.G464S variant had a history of self-mutilation, osteomyelitis, and sensory neuropathy; congenital insensitivity to pain was reported in Individual 3, but nerve conduction studies were not performed. Individual 16 also engaged in self-injurious behavior including tongue and lip biting.

The most profoundly affected individuals exhibited considerable phenotypic overlap with *Flvcr1* knockout mice. Individuals 9-11 in Family 8 were stillborn and exhibited craniofacial, limb, and digital malformations; they were recently identified in a large cohort that investigated the utility of long-read whole genome sequencing^26^ (**Fig. 2X-g**, **Supplemental Data**). Individual 17 from Family 12 died on in the neonatal period. Fetal MRI and autopsy demonstrated craniofacial, limb, and digital malformations in addition to congenital heart disease, renal agenesis, hepatosplenomegaly, and paper-thin cerebral cortex with hydrocephalus and multiple hemorrhages (**Fig. 2X-d**, **Supplemental Data**). Bone marrow hematopoiesis was normal.

Hepatomegaly and milder digital malformations including polydactyly and arthrogryposis were also observed in other individuals with *FLVCR1*-associated severe developmental disorders. Unexplained macrocytic anemia was present in 5 of 12 individuals who survived the neonatal period and for whom red blood cell studies were performed. Reduced fetal movements were reported in Families 8 and 11.

Families 13-20 had mild *FLVCR1*-related diseases including typical phenotypes (PCARP, HSAN, RP) as well as developmental delay and the novel phenotype hereditary spastic paraplegia. While spasticity is a common feature of severe *FLVCR1*-related developmental disorders, it is uncommon in mild *FLVCR1*-related disease^27^. Two recurrent variants, p.Y128N and p.I343T, were observed in unrelated Persian families. Brain MRI was not performed in most instances of mild *FLVCR1*-related disease but was normal in individual 20 with *FLVCR1*-related sensory neuropathy, RP, and mild intellectual disability. The two siblings in Family 17 were evaluated for developmental delay, microcephaly, hypotonia, and hyporreflexia.

There was intrafamilial variability with the younger brother exhibiting more developmental impairment than the older brother (**Supplemental Fig. 2**). In Family 18, the young adult asymptomatic sister of Individual 25 was also homozygous for *FLVCR1*: c.502C>G p.L168V; as age of onset as late as the third or fourth decade of life has been described, this likely represents age-related penetrance^28^.

Twenty *FLVCR1* variants were identified within the cohort; only two, c.1593+5_1593+8del and c.574T>C p.C192R were previously reported (**Table 2**)^19,29^. *FLVCR1*(NM_014053.4) consists of 10 exons and encodes a protein with 12 transmembrane domains. We visualize the distribution of reported pathogenic *FLVCR1* variants within the *FLVCR1*(NM_014053.4) cDNA and protein (**Fig. 3A,B**). All pathogenic *FLVCR1* variants are rare and absent in the homozygous state in gnomAD v2.1.1^30^. Most are predicted damaging by multiple pathogenicity predictors and affect conserved residues (**Table 2**, **Supplemental Table 1**). Amino acid substitutions within FLVCR1 transmembrane domains are predicted damaging by the protein language model ESM1b^31^, and nearly all pathogenic *FLVCR1* missense variants fall within well-conserved transmembrane domains (**Fig. 3B; Supplemental Fig. 4**). All *FLVCR1* nonsense and frameshift variants are predicted to trigger nonsense-mediated decay (NMD)^32^.

**Table 2:**
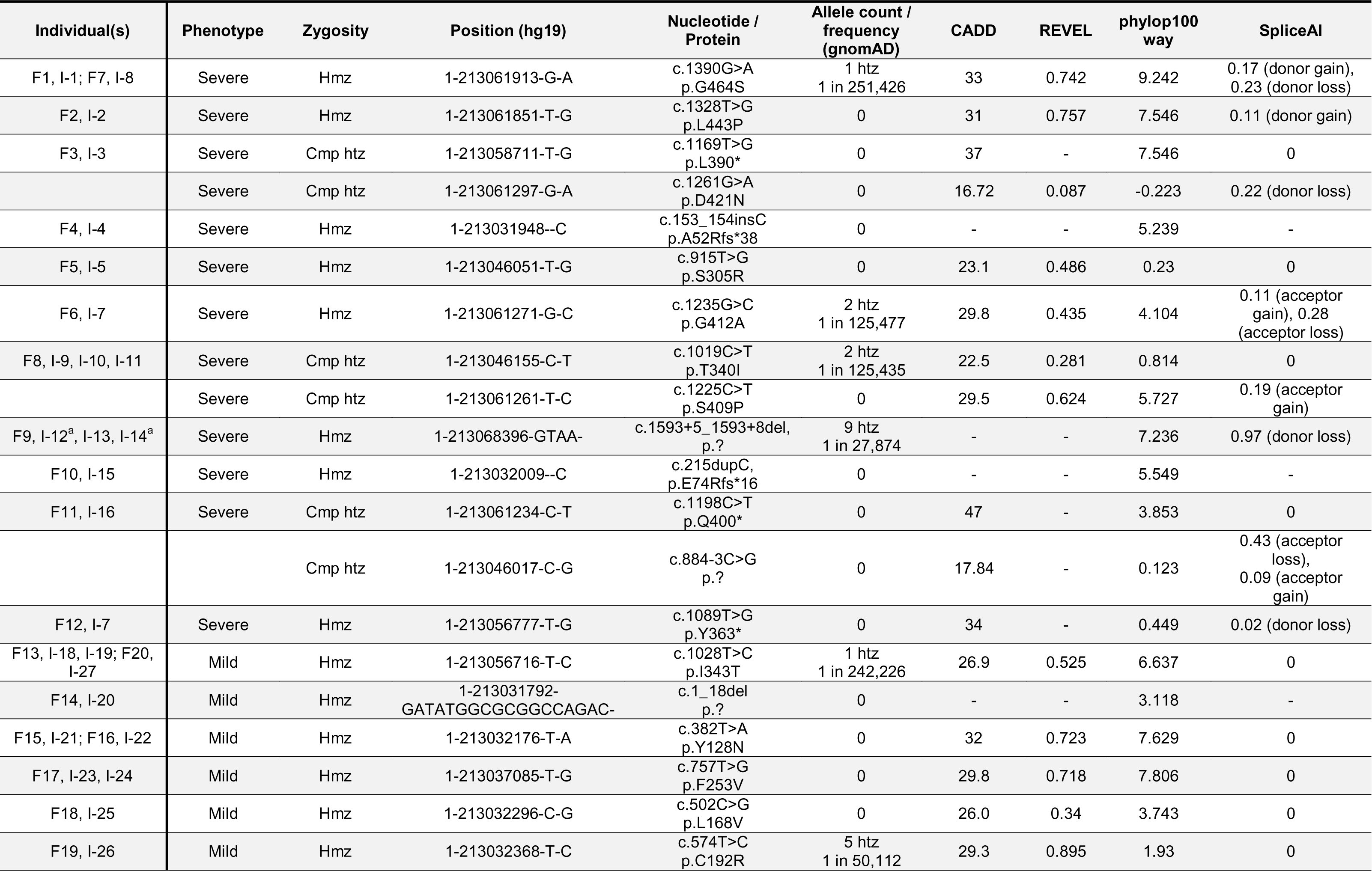
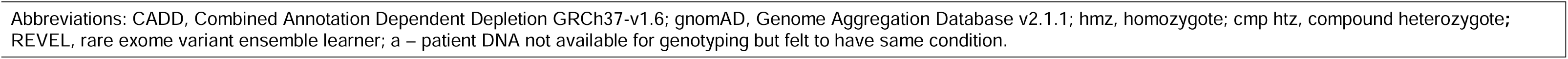
Summary of *FLVCR1* variant alleles

**Figure 3:**
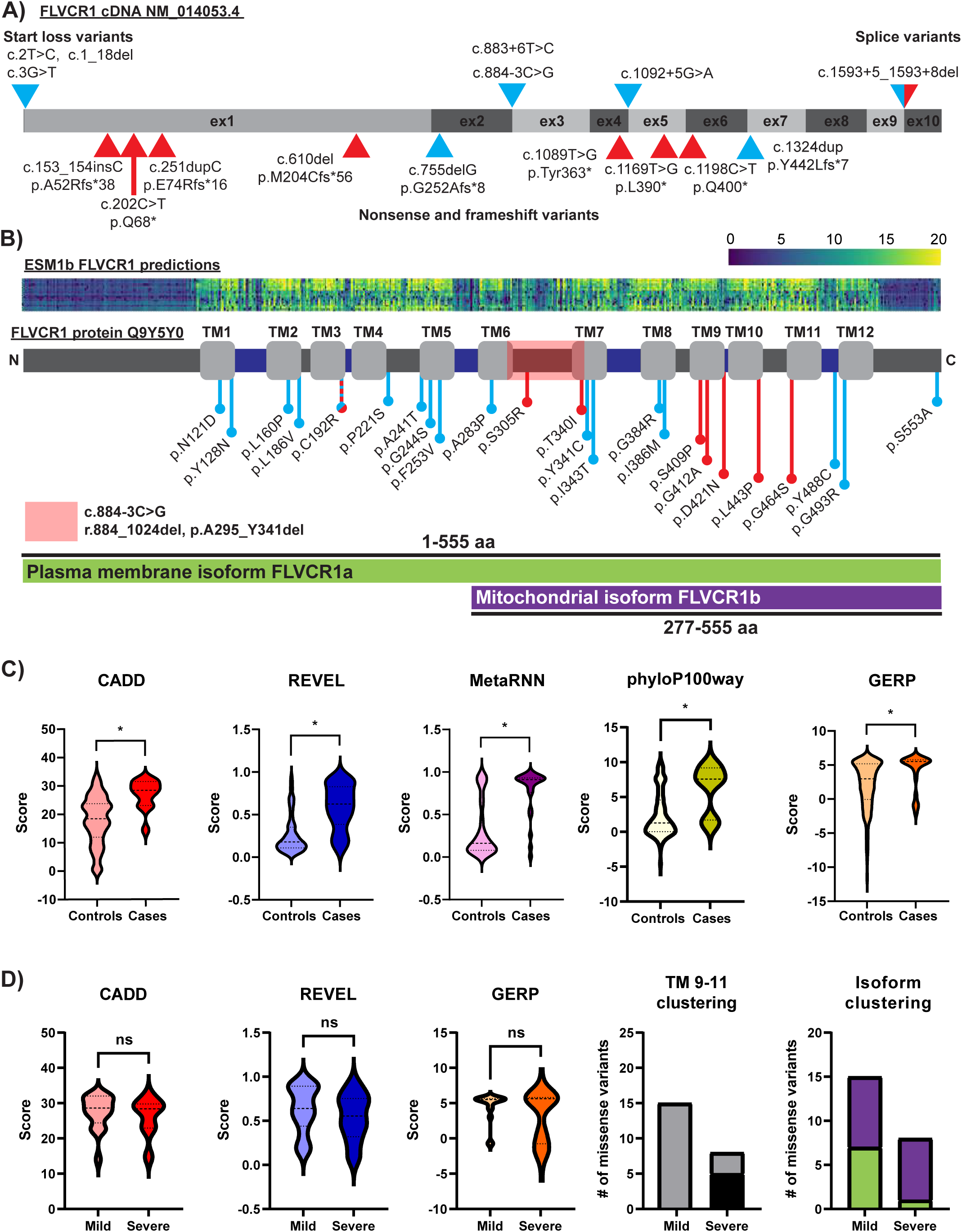
Summary of the *FLVCR1* allelic series. **A)** Model of protein-coding portions of human *FLVCR1* cDNA (NM_014053.4) showing location of novel and previously published start loss variants, splicing variants, nonsense variants, and frameshift variant alleles. Start loss and splicing variants are indicated by arrowheads above the model. Nonsense and frameshift variants are indicated by arrowheads below the model. Red arrowheads show variants associated with severe phenotypes, whereas light blue shading indicates the relatively milder neurological disease. **B)** Model of human FLVCR1 protein (Uniprot entry Q9Y5Y0) showing location of novel and previously published missense variants. Light gray rectangles indicate transmembrane domains, dark gray rectangles indicate intracellular domains, and dark blue rectangles indicate extracellular domains. Red ball and sticks indicate severe phenotypes, whereas light blue ball and sticks indicate mild phenotypes. Portions of FLVCR1 found within the plasma membrane isoform FLVCR1a and mitochondrial isoform FLVCR1b are indicated below the figure in green and purple, respectively. Red box indicates region deleted due to c.884-3C>G (r.884_1024del, p.A295_Y341del). ESM1b pathogenicity predictions are shown above the model. Increasing likelihood of pathogenicity is reflected by the blue (low) to yellow (high) gradient. **C)** Comparison of pathogenicity predictions (CADD, REVEL, MetaRNN) and conservation metrics (phyloP100way, GERP) between novel and previously reported pathogenic *FLVCR1* variants and all variants within gnomAD v3.1.2 (76,156 human samples, 189 variants) and primateseq v1.0 (811 primate samples from 236 primate species, 209 variants). * p<0.05, unpaired t test with Welch’s correction. **D)** Comparison of pathogenicity predictions (CADD, REVEL) and conservation metric (GERP) between mild and severe *FLVCR1*-related phenotypes. Differences in transmembrane domains 9-11 and isoform clustering of *FLVCR1* variants between mild and severe phenotypes is also displayed. Gray = variants located outside transmembrane domains 9-11, black = variants located inside transmembrane domains 9-11, purple = variants located inside mitochondrial isoform FLVCR1b, green = variants located inside plasma membrane isoform FLVCR1a but not FLVCR1b. ns = not significant, TM = transmembrane.

To assess the impact of c.884-3C>G on splicing, whole blood RNA was isolated from the carrier mother of Individual 16, and reverse transcriptase polymerase chain reaction (RT-PCR) was performed. RT-PCR produced two bands: a larger band seen in controls and a smaller band absent in controls (**Supplemental Fig. 4**). Sanger sequencing confirmed that the larger band represented wild-type *FLVCR1*, whereas the smaller band represented a heterozygous deletion of exon 3 (r.884_1024del, p.A295_Y341del). The deletion spans portions of transmembrane domains 6 and 7 and completely removes the fourth cytoplasmic loop (**Fig. 3B**). The splicing variant c.1593+5_1593+8del is strongly predicted to alter splicing by SpliceAI (donor loss Δ score 0.97) and Pangolin (splice loss Δ score 0.88). A mini-gene splicing assay confirmed this variant causes exon 9 skipping which is predicted to result in protein truncation (r.1526_1593del; p.A509Dfs*4) (**Supplemental Fig. 5 & 9**).

Pathogenic missense variants are anticipated to disrupt gene function and involve more highly conserved functional regions than benign missense variants within the human population and non-human primates. We therefore compared pathogenicity predictions and conservation metrics between *FLVCR1* missense variants identified in individuals with *FLVCR1*-related disorders and *FLVCR1* missense variants present in human and primate populations in gnomAD v2.1.1 and primAD v1.0 (**Fig. 3C**)^30,33^. CADD, REVEL, MetaRNN, phyloP100way, and GERP scores were all significantly greater in cases than controls. In contrast, there was no difference in CADD, REVEL, or GERP scores between missense variants identified in mild versus severe *FLVCR1*-related disorders (**Fig. 3D**). Comparison of the distribution of severe versus mild disease-associated missense variants demonstrated that severe disease-associated missense variants form a cluster within transmembrane domains 9-11 (5 of 8 severe disease-associated variants vs. 0 of 15 mild disease-associated variants). Similarly, 87.5% of severe disease-associated missense variants (7/8) lie within both the plasma membrane isoform FLVCR1a and the mitochondrial isoform FLVCR1b, whereas only 53.3% of mild disease-associated missense variants (8/15) fell within both isoforms.

To investigate the molecular consequence of *FLVCR1* missense variants on gene function, we generated missense variants using site-directed mutagenesis of human *FLVCR1* cDNA and examined each variant’s choline and ethanolamine transport activity (**Fig. 4**)^14^.

**Figure 4:**
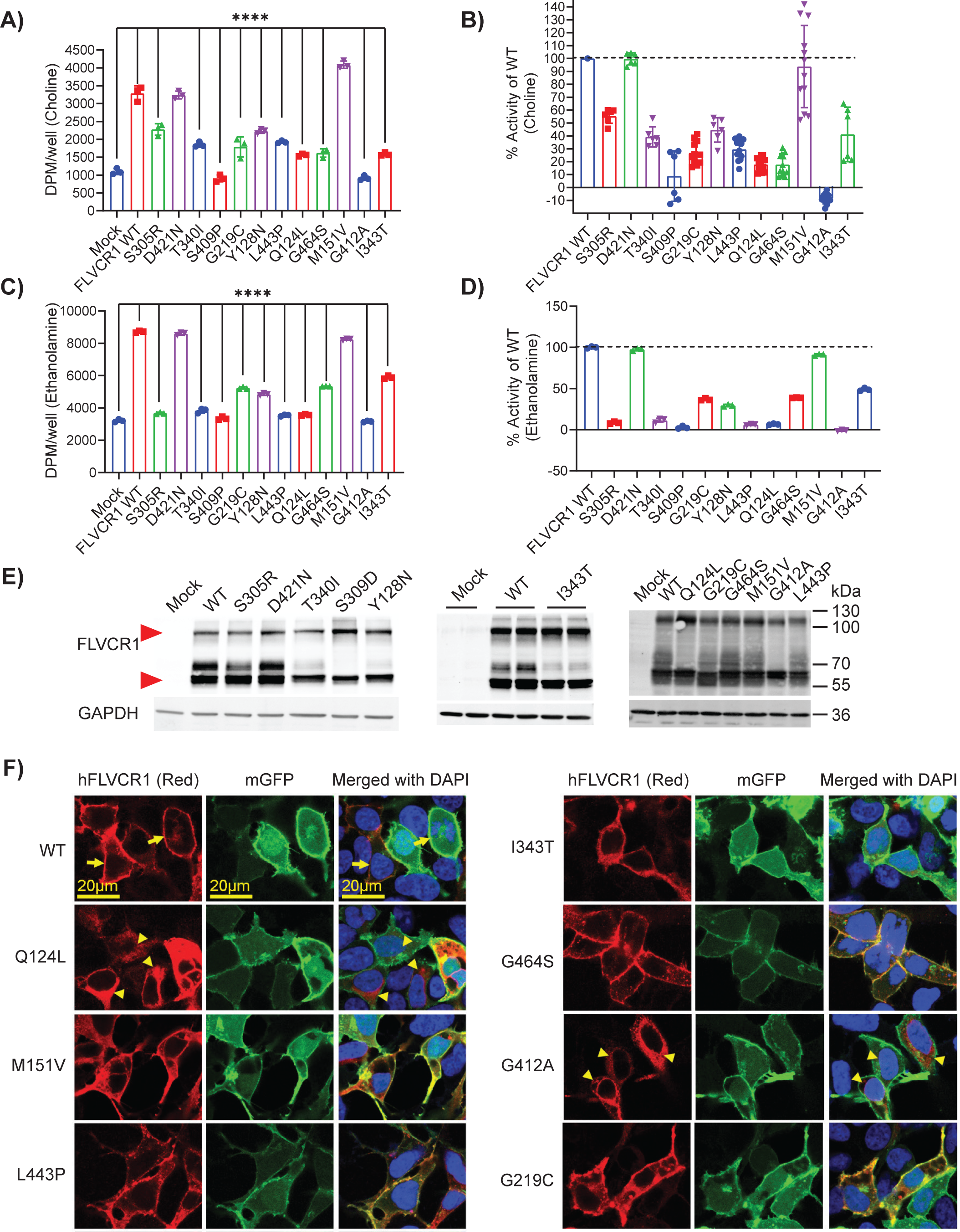
Pathogenic missense variation in *FLVCR1* reduces choline and ethanolamine transport. **A-B)** Choline transport activity of the FLVCR1 missense variants expressed as absolute values (DPM/well) or as percentage of wildtype (WT) FLVCR1. These missense variants and WT human FLVCR1 (hFLVCR1) were co-expressed with choline kinase alpha (CHKA) in HEK293 cells for activity assay with [3H] choline. Mock was transfected with CHKA alone. Experiments were performed at least twice in triplicates. Data are expressed as mean ± SD. ****P<0.0001; One-way ANOVA for A-B. ns, not significant. **C-D)** Ethanolamine transport activity of the FLVCR1 missense variants expressed as absolute values (DPM/well) or as percentage of WT FLVCR1. These missense variants and WT human FLVCR1 were co-expressed with ethanolamine kinase 1 (ETNK1) in HEK293 cells for activity assay with [14C] ethanolamine. Mock was transfected with ETNK1 alone. Experiments were performed at least twice in triplicates. Data are expressed as mean ± SD. ****P<0.0001; One-way ANOVA for C-D. ns, not significant. **E)** Western blot analysis of HEK293 cells overexpressed with WT or hFLVCR1 variants. Red vertical arrowhead marks M_r_ = 59 kD migration of FLVCR1; GAPDH loading control shown below. **F)** Immunostaining of WT or hFLVCR1 mutants overexpressed in HEK293 cells. Vertical yellow horizontal bar shows image scale. Arrows (yellow) show plasma membrane, where membrane GFP was used as a marker. Arrowheads show intracellular signals of hFLVCR1.

Briefly, human *FLVCR1* cDNA was co-transfected into HEK293 cells along with human choline kinase A (*CHKA*) or ethanolamine kinase 1 (*ETNK1*) cDNA because the sole expression of *FLVCR1* cDNA only slightly increased choline or ethanolamine transport activity. HEK293 cells were then incubated with [^3^H] choline or [^14^C] ethanolamine, washed, and cellular radioactivity levels were measured. CHKA catalyzes choline phosphorylation into phosphocholine while ETNK1 catalyzes ethanolamine into phosphatidylethanolamine; their co-expression along with FLVCR1 greatly increases choline or ethanolamine import^14^. In addition to the FLVCR1 missense variants identified in this publication, we also examined p.Q124L and p.G219C, two FLVCR1 variants recently identified in an individual with HSAN^19,34^. These assays demonstrated nearly all FLVCR1 missense variants significantly reduced choline and ethanolamine transport activity relative to wild-type FLVCR1 with transport activity ranging from 0% to 55.38% (choline) and 0% to 48.80%. The transport activity of two variants, p.M151V and p.D421N, was comparable to wild-type FLVCR1. The p.M151V variant occurred in the homozygous state in an individual within the BCM-GREGoR database. This individual had a blended developmental disorder phenotype resulting from multi-locus pathogenic variation in three genes: *AP4B1*, *AMPD2*, and *NOTCH2*^35^. As the individual’s phenotype is explained by other gene variants and the p.M151V variant’s transport activity is normal, p.M151V may represent a rare benign polymorphism. The p.D421N variant was identified in compound heterozygosity with a nonsense variant in Individual 3. Although p.D421N did not alter choline or ethanolamine transport activity, the variant is weakly predicted to cause donor loss by SpliceAI (score 0.22). A mini-gene splicing assay of this variant (c.1261G>A p.D421N) demonstrated a strong effect on exon 6 skipping (**Supplemental Fig. 5 & 6**). The phenotypic overlap of Individual 3 with other severe *FLVCR1* cases and the presence of typical *FLVCR1* features including RP and posterior column T2 hyperintensity supports the variant’s pathogenicity. Another missense variant in the same exon (c.1235G>A p.G412A) also showed a strong effect on exon 6 skipping in addition to its effect on choline and ethanolamine transport (**Supplemental Fig. 5 & 6**). Western blot analysis of HEK293 cells overexpressing wild-type and variant FLVCR1 showed similar levels of FLVCR1 protein indicating that FLVCR1 missense variants do not impact protein stability (**Fig. 4**). Immunostaining demonstrates FLVCR1 missense variants localize to the plasma membrane like wild-type FLVCR1; only p.G412A exhibited abnormal intracellular accumulation (**Fig. 4**). As we previously characterized the choline transport activity of all *FLVCR1* missense variants reported in the literature^36^, we compared transport activity between mild and severe phenotype-associated variants but found no significant difference (mean choline transport activity as a percentage of wild-type FLVCR1: mild phenotypes, 34.3±21.9; severe phenotypes, 35.5±21.9; p=0.9222).

Here, we demonstrate rare genetic variation in the choline and ethanolamine transporter gene *FLVCR1* causes a broad and pleiotropic recessive disease spectrum ranging from adult neurodegeneration to severe developmental disorders. Choline is an essential nutrient which plays an integral role in methyl group metabolism, phosphatidylcholine (PC) synthesis via the Kennedy pathway, and acetylcholine synthesis^37^. While *de novo* choline synthesis occurs in the liver through the phosphatidylethanolamine *N*-methyltransferase pathway, this pathway is inadequate to meet the needs of the human organism^37^. Choline is found in animal products, cruciferous vegetables, and beans^37^. The dietary intake of choline in most individuals in the United States and Europe falls below the United States Institute of Medicine’s adequate intake level^37,38^. Choline insufficiency is particularly common in low-income countries, and choline is neglected in nutrient-fortified food aid^39^. Furthermore, individual dietary requirements vary considerably due to multiple factors including genotype, sex, developmental stage, and dietary intake of vitamins involved in methyl group metabolism including folate and B12^40^. Single nucleotide polymorphisms in genes involved in the phosphatidylethanolamine *N*-methyltransferase pathway, folate and methyl group metabolism, and choline transport including *PEMT*, *MTHFR*, *MTR*, *MTRR*, and *SLC44A1* have been shown to modulate choline requirements and risk of choline deficiency^40^. Similarly, ethanolamine cannot be synthesized in humans and is a precursor for phosphatidylethanolamine (PE) synthesis via the Kennedy pathway^41^. PE and PC are abundant membrane phospholipids required for membrane integrity, cell division, and mitochondrial respiratory function.

Choline is required for normal neurodevelopment^42^. Maternal choline deficiency impairs hippocampal development as well as neuronal progenitor cell and retinal progenitor cell expansion and differentiation in mouse embryos^43,44^. Choline deficiency also causes anemia, liver disease, growth retardation, and immune deficiency^45–47^. Neurodevelopment is also disrupted by defective choline uptake^48^. Pathogenic variation in *SLC5A7*, the gene encoding a high affinity choline transporter expressed in cholinergic neurons, causes two neurodevelopmental disorders: autosomal recessive (AR) presynaptic congenital myasthenic syndrome 20 [MIM: 617143] and autosomal dominant (AD) distal hereditary motor neuronopathy [MIM: 158580]^49,50^. Similarly, pathogenic variation in *SLC44A1*, a gene predicted to encode the low affinity choline transporter CTL1, causes AR childhood-onset neurodegeneration with ataxia, tremor, optic atrophy, and cognitive decline [MIM: 618868]^51^.

Finally, the protein encoded by closely related gene *FLVCR2* is expressed at the blood-brain barrier and transports choline and ethanolamine^16,52^. Pathogenic variation in *FLVCR2* causes Fowler syndrome, a severe AR disorder characterized by proliferative vasculopathy, hydranencephaly, fetal akinesia deformation sequence, and prenatal lethality [MIM: 225790]^53^. The identification of *FLVCR1* as a neurodevelopmental choline and ethanolamine transport disorder further reinforces the critical role of choline and ethanolamine in brain development. Additionally, the identification of multiple neurodevelopmental choline and ethanolamine transport disorders demonstrates considerable complexity to choline and ethanolamine regulation within the developing brain and warrants further study. Therapeutic choline and ethanolamine supplementation has not been attempted in patients with choline and ethanolamine transport disorders but could in theory be efficacious as in riboflavin transporter deficiency^6^. The existence of multiple choline transporters allows for alternative routes for choline uptake^1^, and supplementation could potentially boost choline intake through hypomorphic transporters. Randomized controlled trials of choline supplementation in fetal alcohol syndrome have shown choline supplementation is well-tolerated and can have beneficial neurocognitive effects^54,55^. Measurement of choline and ethanolamine levels in biospecimens from individuals with *FLVCR1*-related disorders is needed to inform choline and/or ethanolamine supplementation as a potential therapeutic modality.

Based on these findings, we propose an allele-specific gene dosage model in which disease severity is a function of residual FLVCR1 activity. This proposal stems from three key observations. First, we identified three individuals with severe developmental disorders and homozygous *FLVCR1* pLoF variants. These individuals are thus genetically equivalent to *Flvcr1* knockout mice and zebrafish, which also exhibit severe developmental phenotypes^11,56^. Biallelic pLoF *FLVCR1* variants have not been identified in individuals with milder *FLVCR1*-related phenotypes like PCARP; nearly all reported individuals instead have biallelic missense variants. Second, we show most pathogenic *FLVCR1* missense variants reduce but do not completely eliminate choline and ethanolamine transport and thus likely represent hypomorphic alleles^36^.

*Flvcr1* was previously shown to exhibit highest central nervous system expression in the retina followed by the posterior column of the spinal cord and cerebellum; this may parsimoniously explain the susceptibility of these tissues to mild hypomorphic alleles^19^. Third, we observe evidence of complex compound inheritance in which *FLVCR1* variants may contribute to mild phenotypes in homozygosity but severe phenotypes in compound heterozygosity with a more severe variant (e.g., *FLVCR1* p.C192R) or severe phenotypes in homozygosity but mild phenotypes when paired with a less deleterious variant (e.g., *FLVCR1* c.1593+5_1593+8del)^19,21,29^. The allele-specific gene dosage model is well-described in recessive disorders but can confound diagnostic personalized genome analysis when the clinically reported phenotype diverges substantially from the disease trait clinical synopsis provided in the literature or online databases like Online Mendelian Inheritance in Man (OMIM). Indeed, the severely affected individuals reported here had all undergone clinical or research exome or genome sequencing which identified the reported *FLVCR1* variants, yet in each case the variants were previously felt either non-contributory or of uncertain significance given the apparent phenotypic mismatch. Such false assumptions illustrate the importance of incorporating model organism data into personalized genome analysis for rare diseases and the need to anticipate more severe and milder phenotypes associated with each disease gene locus to maximize the yield of diagnostic genetic testing.

We did not observe a correlation between FLVCR1 missense variant choline transport activity and phenotypic severity in our functional assay. There are several possible explanations. First, our choline transport assay may lack the sensitivity to discriminate between mild and severe phenotype-associated variants. As the transport assay is reliant on overexpression of *FLVCR1* cDNA, it is insensitive to splicing defects. Indeed, we demonstrate through mini-gene splicing assays that several *FLVCR1* single nucleotide variants and indels disrupt splicing (**Supplemental Figures 5-9**). Second, the cellular phenotype assessed by the choline transport assay must be distinguished from the organismal phenotypes characterized in humans with pathogenic *FLVCR1* variation. Third, FLVCR1 was found to transport additional ligands like ethanolamine in both this study and previous studies^14^. While a reduction in both choline and ethanolamine transport was observed with most variants, some like p.S305R had a more severe impact on ethanolamine than choline transport. Fourth, as both plasma membrane and mitochondrial isoforms FLVCR1a and FLVCR1b are required for murine viability^9^, a missense variant involving both isoforms may have a greater phenotypic impact than one only impacting the plasma membrane isoform FLVCR1a. In line with this hypothesis, we observed nearly all severe disease associated *FLVCR1* missense variants are predicted to impact both isoforms. Another possibility is maternal and/or fetal deficiency of choline, ethanolamine, folate, and/or B12 due to poor dietary intake or maternal-fetal genotypes could act as phenotypic modifiers^40^. Finally, it is important to note the phenotypes associated with homozygous first exon frameshifts in Individuals 4 and 15 were not as severe as those seen in Individual 17 with the homozygous exon 4 nonsense variant nor in individuals 12-14 with the homozygous splice site variant c.1593+5_1593+8del. This defines a gene polarity effect^57^. One possible explanation is first exon pLoF variants may not represent true null alleles. First exon premature termination codons (PTCs) can escape NMD and generate a functional or partially functional truncated protein if a downstream methionine allows for in-frame translation re-initiation^58^.

In summary, our report reveals a broad and pleiotropic phenotypic spectrum resulting from biallelic *FLVCR1* variants; these range from adult neurodegeneration to severe developmental disorders with variable anemia and skeletal malformations. Combined with recent studies demonstrating *FLVCR1* encodes a choline and ethanolamine transporter^4,13,16,36^, these data suggest choline and ethanolamine transport into the central and peripheral nervous systems is essential to prevent neurodegeneration and required for neurodevelopment. The observation that the most severe *FLVCR1*-related phenotypes cause stillbirth and resemble Diamond-Blackfan anemia establishes *FLVCR1* should be considered in the differential diagnosis of recurrent miscarriage, multiple congenital anomalies, and severe Diamond-Blackfan anemia-like phenotypes. Further studies are required to understand the impact of gene x environmental (GxE) interactions on *FLVCR1*-related phenotypes and the therapeutic implications of choline or ethanolamine supplementation in *FLVCR1*-related diseases.

## Web Resources

Online Mendelian Inheritance in Man, http://www.omim.org

gnomAD Browser, https://gnomad.broadinstitute.org/

Baylor College of Medicine Human Genome Sequencing Center, https://www.hgsc.bcm.edu

Baylor College of Medicine Lupski Lab, https://github.com/BCM-Lupskilab

CADD, https://cadd.gs.washington.edu/

SpliceAI and Pangolin, https://spliceailookup.broadinstitute.org/

primAD Browser, https://primad.basespace.illumina.com/

## Data Availability

All data described in this study are provided within the article and Supplementary Material. Raw sequencing data and de-identified clinical data are available from the corresponding authors upon request.

## Data Availability

All data produced in the present study are available upon reasonable request to the authors.

## Acknowledgements

This study was supported in part by the U.S. National Human Genome Research Institute (NHGRI) and National Heart Lung and Blood Institute (NHBLI) to the Baylor-Hopkins Center for Mendelian Genomics (BHCMG, UM1 HG006542, J.R.L); NHGRI grant as part of the GREGoR Consortium (U01 HG011758 to J.E.P., J.R.L., and R.A.G.); NHGRI grant to Baylor College of Medicine Human Genome Sequencing Center (U54HG003273 to R.A.G.); U.S. National Institute of Neurological Disorders and Stroke (NINDS) (R35NS105078 to J.R.L); Muscular Dystrophy Association (MDA) (512848 to J.R.L.); Singapore Ministry of Education grants (T2EP30221-0012, T2EP30123-0014, and NUHSRO/2022/067/T1 to L.N.N.); and Spastic Paraplegia Foundation Research Grant to J.R.L. D.P. was supported by a NINDS 1K23 NS125126-01A1 and Rett Syndrome Research Trust fellowship award from International Rett Syndrome Foundation (IRSF grant #3701-1). D.G.C. was supported by NIH Medical Genetics Research Fellowship Program (T32 GM007526), the Chao Physician Scientist Award, the Child Neurologist Career Development Program K12, and MDA Development Grant (873841). K.M.B was supported by the GREGoR Consortium Research Grant from the GREGoR Data Coordinating Center (U24HG011746 to KMB); Foundation Fighting Blindness (EGI-GE-1218-0753-UCSD to KMB), Iraty Award 2023, Lions Foundation, Research to Prevent Blindness, and NEI: P30EY014104 (MEEI core support).

## Ethics Declaration

This study adheres to the principles in the Declaration of Helsinki. The study was approved by Baylor College of Medicine Institutional Review Board (IRB) protocol H-29697. Informed consent including consent to have the results of this research work published was obtained from all participants as required by the IRB.

## References

1. Pizzagalli MD, Bensimon A, Superti-Furga G. A guide to plasma membrane solute carrier proteins. FEBS J. 2021;288(9):2784–2835. doi:10.1111/febs.15531

2. Marafi D, Fatih JM, Kaiyrzhanov R, et al. Biallelic variants in SLC38A3 encoding a glutamine transporter cause epileptic encephalopathy. Brain. 2022;145(3):909–924. doi:10.1093/brain/awab369

3. Saida K, Maroofian R, Sengoku T, et al. Brain monoamine vesicular transport disease caused by homozygous SLC18A2 variants: A study in 42 affected individuals. Genet Med. 2023;25(1):90–102. doi:10.1016/j.gim.2022.09.010

4. Kenny TC, Khan A, Son Y, et al. Integrative genetic analysis identifies FLVCR1 as a plasma-membrane choline transporter in mammals. Cell Metab. 2023;35(6):1057–1071.e12. doi:10.1016/j.cmet.2023.04.003

5. Klepper J, Akman C, Armeno M, et al. Glut1 Deficiency Syndrome (Glut1DS): State of the art in 2020 and recommendations of the international Glut1DS study group. Epilepsia Open. 2020;5(3):354–365. doi:10.1002/epi4.12414

6. O’Callaghan B, Bosch AM, Houlden H. An update on the genetics, clinical presentation, and pathomechanisms of human riboflavin transporter deficiency. J Inherit Metab Dis. 2019;42(4):598–607. doi:10.1002/jimd.12053

7. Mathiesen BK, Miyakoshi LM, Cederroth CR, et al. Delivery of gene therapy through a cerebrospinal fluid conduit to rescue hearing in adult mice. Sci Transl Med. 2023;15(702):eabq3916. doi:10.1126/scitranslmed.abq3916

8. Tailor CS, Willett BJ, Kabat D. A putative cell surface receptor for anemia-inducing feline leukemia virus subgroup C is a member of a transporter superfamily. J Virol. 1999;73(8):6500–6505. doi:10.1128/JVI.73.8.6500-6505.1999

9. Chiabrando D, Marro S, Mercurio S, et al. The mitochondrial heme exporter FLVCR1b mediates erythroid differentiation. J Clin Invest. 2012;122(12):4569–4579. doi:10.1172/JCI62422

10. Quigley JG, Yang Z, Worthington MT, et al. Identification of a human heme exporter that is essential for erythropoiesis. Cell. 2004;118(6):757–766. doi:10.1016/j.cell.2004.08.014

11. Keel SB, Doty RT, Yang Z, et al. A heme export protein is required for red blood cell differentiation and iron homeostasis. Science. 2008;319(5864):825–828. doi:10.1126/science.1151133

12. Ponka P, Sheftel AD, English AM, Scott Bohle D, Garcia-Santos D. Do Mammalian Cells Really Need to Export and Import Heme? Trends Biochem Sci. 2017;42(5):395–406. doi:10.1016/j.tibs.2017.01.006

13. Tsuchiya M, Tachibana N, Nagao K, Tamura T, Hamachi I. Organelle-selective click labeling coupled with flow cytometry allows high-throughput CRISPR screening of genes involved in phosphatidylcholine metabolism. Published online April 18, 2022:2022.04.18.488621. doi:10.1101/2022.04.18.488621

14. Ha HTT, Sukumar VK, Chua JWB, et al. Mfsd7b facilitates choline transport and missense mutations affect choline transport function. Cell Mol Life Sci. 2023;81(1):3. doi:10.1007/s00018-023-05048-4

15. Son Y, Kenny TC, Khan A, Birsoy K, Hite RK. Structural basis of lipid head group entry to the Kennedy pathway by FLVCR1. BioRxiv Prepr Serv Biol. Published online September 28, 2023:2023.09.28.560019. doi:10.1101/2023.09.28.560019

16. Ri K, Weng TH, Cabezudo AC, et al. Structural and mechanistic insights into human choline and ethanolamine transport. Published online December 19, 2023:2023.09.15.557925. doi:10.1101/2023.09.15.557925

17. Da Costa L, Leblanc T, Mohandas N. Diamond-Blackfan anemia. Blood. 2020;136(11):1262-1273. doi:10.1182/blood.2019000947

18. Rey MA, Duffy SP, Brown JK, et al. Enhanced alternative splicing of the FLVCR1 gene in Diamond Blackfan anemia disrupts FLVCR1 expression and function that are critical for erythropoiesis. Haematologica. 2008;93(11):1617–1626. doi:10.3324/haematol.13359

19. Rajadhyaksha AM, Elemento O, Puffenberger EG, et al. Mutations in FLVCR1 cause posterior column ataxia and retinitis pigmentosa. Am J Hum Genet. 2010;87(5):643–654. doi:10.1016/j.ajhg.2010.10.013

20. Kuehlewein L, Schöls L, Llavona P, et al. Phenotypic spectrum of autosomal recessive retinitis pigmentosa without posterior column ataxia caused by mutations in the FLVCR1 gene. Graefes Arch Clin Exp Ophthalmol. 2019;257(3):629–638. doi:10.1007/s00417-018-04233-7

21. Chiabrando D, Castori M, di Rocco M, et al. Mutations in the Heme Exporter FLVCR1 Cause Sensory Neurodegeneration with Loss of Pain Perception. PLoS Genet. 2016;12(12):e1006461. doi:10.1371/journal.pgen.1006461

22. Calame DG, Guo T, Wang C, et al. Monoallelic variation in DHX9, the gene encoding the DExH-box helicase DHX9, underlies neurodevelopment disorders and Charcot-Marie-Tooth disease. Am J Hum Genet. 2023;110(8):1394–1413. doi:10.1016/j.ajhg.2023.06.013

23. Sobreira N, Schiettecatte F, Valle D, Hamosh A. GeneMatcher: a matching tool for connecting investigators with an interest in the same gene. Hum Mutat. 2015;36(10):928–930. doi:10.1002/humu.22844

24. Wohler E, Martin R, Griffith S, et al. PhenoDB, GeneMatcher and VariantMatcher, tools for analysis and sharing of sequence data. Orphanet J Rare Dis. 2021;16(1):365. doi:10.1186/s13023-021-01916-z

25. Coban-Akdemir Z, Song X, Ceballos FC, et al. De novo mutation and identity-by-descent drive disease haplotypes, biallelic traits and multilocus pathogenic variation. Published online July 27, 2022:2020.04.27.064824. doi:10.1101/2020.04.27.064824

26. AlAbdi L, Shamseldin HE, Khouj E, et al. Beyond the exome: utility of long-read whole genome sequencing in exome-negative autosomal recessive diseases. Genome Med. 2023;15(1):114. doi:10.1186/s13073-023-01270-8

27. Vaughan DP, Costello DJ. Extending the phenotype of posterior column ataxia with retinitis pigmentosa caused by variants in FLVCR1. Am J Med Genet A. 2022;188(4):1259–1262. doi:10.1002/ajmg.a.62612

28. Li Z, Li Y, Chu X, et al. Novel mutations in FLVCR1 cause tremors, sensory neuropathy with retinitis pigmentosa. Neuropathol Off J Jpn Soc Neuropathol. Published online July 19, 2023. doi:10.1111/neup.12936

29. Shaibani A, Wong LJ, Wei Zhang V, Lewis RA, Shinawi M. Autosomal recessive posterior column ataxia with retinitis pigmentosa caused by novel mutations in the FLVCR1 gene. Int J Neurosci. 2015;125(1):43–49. doi:10.3109/00207454.2014.904858

30. Karczewski KJ, Francioli LC, Tiao G, et al. The mutational constraint spectrum quantified from variation in 141,456 humans. Nature. 2020;581(7809):434–443. doi:10.1038/s41586-020-2308-7

31. Brandes N, Goldman G, Wang CH, Ye CJ, Ntranos V. Genome-wide prediction of disease variant effects with a deep protein language model. Nat Genet. 2023;55(9):1512–1522. doi:10.1038/s41588-023-01465-0

32. Coban-Akdemir Z, White JJ, Song X, et al. Identifying Genes Whose Mutant Transcripts Cause Dominant Disease Traits by Potential Gain-of-Function Alleles. Am J Hum Genet. 2018;103(2):171–187. doi:10.1016/j.ajhg.2018.06.009

33. Gao H, Hamp T, Ede J, et al. The landscape of tolerated genetic variation in humans and primates. Science. 2023;380(6648):eabn8153. doi:10.1126/science.abn8197

34. Suthar R, Sharawat IK, Eggermann K, et al. Hereditary Sensory and Autonomic Neuropathy: A Case Series of Six Children. Neurol India. 2022;70(1):231. doi:10.4103/0028-3886.338691

35. Karaca E, Posey JE, Akdemir ZC, et al. Phenotypic expansion illuminates multilocus pathogenic variation. Genet Med. 2018;20(12):1528–1537. doi:10.1038/gim.2018.33

36. Ha HTT, Sukumar VK, Chua JWB, et al. Mfsd7b facilitates choline uptake and missense mutations affect choline transport function. Published online September 30, 2023:2023.09.30.560304. doi:10.1101/2023.09.30.560304

37. Zeisel SH, Klatt KC, Caudill MA. Choline. Adv Nutr. 2018;9(1):58-60. doi:10.1093/advances/nmx004

38. Vennemann FBC, Ioannidou S, Valsta LM, et al. Dietary intake and food sources of choline in European populations. Br J Nutr. 2015;114(12):2046–2055. doi:10.1017/S0007114515003700

39. May T, Caudill M, Manary M. Is There Enough Choline for Children in Food Aid? JAMA Pediatr. 2023;177(3):223–224. doi:10.1001/jamapediatrics.2022.5543

40. Ganz AB, Klatt KC, Caudill MA. Common Genetic Variants Alter Metabolism and Influence Dietary Choline Requirements. Nutrients. 2017;9(8):837. doi:10.3390/nu9080837

41. Patel D, Witt SN. Ethanolamine and Phosphatidylethanolamine: Partners in Health and Disease. Oxid Med Cell Longev. 2017;2017:4829180. doi:10.1155/2017/4829180

42. Zeisel SH, Niculescu MD. Perinatal choline influences brain structure and function. Nutr Rev. 2006;64(4):197–203. doi:10.1111/j.1753-4887.2006.tb00202.x

43. Trujillo-Gonzalez I, Friday WB, Munson CA, et al. Low availability of choline in utero disrupts development and function of the retina. FASEB J. 2019;33(8):9194–9209. doi:10.1096/fj.201900444R

44. Wang Y, Surzenko N, Friday WB, Zeisel SH. Maternal dietary intake of choline in mice regulates development of the cerebral cortex in the offspring. FASEB J. 2016;30(4):1566–1578. doi:10.1096/fj.15-282426

45. Alexander HD, Engel RW. The importance of choline in the prevention of nutritional edema in rats fed low-protein diets. J Nutr. 1952;47(3):361–374. doi:10.1093/jn/47.3.361

46. Zeisel SH, Da Costa KA, Franklin PD, et al. Choline, an essential nutrient for humans. FASEB J Off Publ Fed Am Soc Exp Biol. 1991;5(7):2093–2098.

47. May T, de la Haye B, Nord G, et al. One-carbon metabolism in children with marasmus and kwashiorkor. EBioMedicine. 2022;75:103791. doi:10.1016/j.ebiom.2021.103791

48. Wortmann SB, Mayr JA. Choline-related-inherited metabolic diseases—A mini review. J Inherit Metab Dis. 2019;42(2):237–242. doi:10.1002/jimd.12011

49. Barwick KES, Wright J, Al-Turki S, et al. Defective presynaptic choline transport underlies hereditary motor neuropathy. Am J Hum Genet. 2012;91(6):1103–1107. doi:10.1016/j.ajhg.2012.09.019

50. Bauché S, O’Regan S, Azuma Y, et al. Impaired Presynaptic High-Affinity Choline Transporter Causes a Congenital Myasthenic Syndrome with Episodic Apnea. Am J Hum Genet. 2016;99(3):753–761. doi:10.1016/j.ajhg.2016.06.033

51. Fagerberg CR, Taylor A, Distelmaier F, et al. Choline transporter-like 1 deficiency causes a new type of childhood-onset neurodegeneration. Brain J Neurol. 2020;143(1):94–111. doi:10.1093/brain/awz376

52. Nguyen XTA, Le TNU, Ha HTT, et al. MFSD7c functions as a transporter of choline at the blood-brain barrier. Published online October 3, 2023:2023.10.03.560597. doi:10.1101/2023.10.03.560597

53. Meyer E, Ricketts C, Morgan NV, et al. Mutations in FLVCR2 are associated with proliferative vasculopathy and hydranencephaly-hydrocephaly syndrome (Fowler syndrome). Am J Hum Genet. 2010;86(3):471–478. doi:10.1016/j.ajhg.2010.02.004

54. Wozniak JR, Fuglestad AJ, Eckerle JK, et al. Choline supplementation in children with fetal alcohol spectrum disorders: a randomized, double-blind, placebo-controlled trial. Am J Clin Nutr. 2015;102(5):1113–1125. doi:10.3945/ajcn.114.099168

55. Gimbel BA, Anthony ME, Ernst AM, et al. Long-term follow-up of a randomized controlled trial of choline for neurodevelopment in fetal alcohol spectrum disorder: corpus callosum white matter microstructure and neurocognitive outcomes. J Neurodev Disord. 2022;14(1):59. doi:10.1186/s11689-022-09470-w

56. Mercurio S, Petrillo S, Chiabrando D, et al. The heme exporter Flvcr1 regulates expansion and differentiation of committed erythroid progenitors by controlling intracellular heme accumulation. Haematologica. 2015;100(6):720–729. doi:10.3324/haematol.2014.114488

57. Inoue K, Khajavi M, Ohyama T, et al. Molecular mechanism for distinct neurological phenotypes conveyed by allelic truncating mutations. Nat Genet. 2004;36(4):361–369. doi:10.1038/ng1322

58. Neu-Yilik G, Amthor B, Gehring NH, et al. Mechanism of escape from nonsense-mediated mRNA decay of human β-globin transcripts with nonsense mutations in the first exon. RNA. 2011;17(5):843–854. doi:10.1261/rna.2401811

