## Supplemental Methods, Figures, and Tables for "Biallelic variation in the choline and ethanolamine transporter *FLVCR1* underlies a pleiotropic disease spectrum from adult neurodegeneration to severe developmental disorders"

**Participant Identification and Recruitment.** This study adheres to the principles in the Declaration of Helsinki. The study was approved by the Baylor College of Medicine (BCM) institutional review board (IRB). All individuals or their guardians provided informed consent under BCM protocol H-29697 or through other collaborative IRBs. Participants were identified either through the Baylor-Hopkins Center for Mendelian Genomics (BHCMG)/BCM Genomics Research Elucidates the Genetics of Rare disease (BCM-GREGoR) database, the Baylor Genetics (BG) clinical diagnostic laboratory database, GeneMatcher^1,2^, or other research and clinical diagnostic laboratories. All individuals were examined by a clinical geneticist and/or neurologist. Pedigrees and deep phenotypic data for all participants were collected from collaborating clinicians using a standardized template**.**

**Exome/Genome Sequencing and Analysis.** Exome sequencing (ES) was performed at Baylor College of Medicine Human Genome Sequencing Center (BCM-HGSC) using an Illumina dual indexed, paired-end pre-capture library per manufacturer protocol with modifications as previously described for Individuals 2 and 6 (https://www.hgsc.bcm.edu/content/protocols-sequencing-library-construction)^3,4^. Libraries were pooled and hybridized to the HGSC VCRome 2.1 plus custom Spike-In design according to the manufacturer’s protocol (NimbleGen) with minor revisions^5^. Paired-end sequencing was performed with the Illumina NovaSeq6000 platform. Samples achieved 98% of the targeted exome bases covered to a depth of 20x or greater and had a sequencing yield of 13.2 Gb. Illumina sequence analysis was performed using the HGSC HgV analysis pipeline which moves data through various analysis tools from the initial sequence generation on the instrument to annotated variant calls (SNPs and intra-read in/dels)^6,7^. In parallel to the exome workflow a SNP Trace panel was generated for a final quality assessment. This included orthogonal confirmation of sample identity and purity using the Error Rate In Sequencing (ERIS) pipeline developed at the BCM-HGSC26. Using an “e-GenoTyping” approach, ERIS screens all sequence reads for exact matches to probe sequences defined by the variant and position of interest. A successfully sequenced sample must meet quality control metrics of ERIS SNP array concordance (>90%) and ERIS average contamination rate (<5%).

Individuals 1 and 17 underwent clinical proband exome sequencing at Baylor Genetics. Individual 3’s clinical trio exome sequencing was performed by GeneDx. Individual 11 underwent trio genome sequencing as part of the Illumina iHope program. Individuals 8 and 16 were sequenced as part of the 100,000 Genomes Project. Individual 13 underwent clinical proband exome sequencing at King Faisal Specialist Hospital and Research Center. Individual 15 underwent trio exome sequencing at Novogene Corporation Inc, as part of a brain malformation cohort collected at Rutgers Robert Wood Johnson Medical School and Alexandria University. The remaining individuals either underwent research ES as part of the Queen Square Genomics (QSG) study or clinical ES through Palindrome.

Rare, predicted damaging (CADD≥15) single nucleotide variants and indel *FLVCR1* variants in the homozygous or compound heterozygous state in individuals without a molecular diagnosis were analysed within the BHCMG/BCM-GREGoR database as well as the BG clinical diagnostic laboratory. Each case was subsequently re-analyzed using a standardized protocol to rule out alternative molecular diagnoses^8^.

All ES data generated by the BHCMG/BCM-GREGoR for which informed consent for deposition into controlled-access databases was provided was deposited into either dbGaP under the BHCMG dbGaP Study Accession phs000711.v7.p2 or in the AnVIL repository under study name Baylor-Hopkins Center for Mendelian Genomics (<https://anvilproject.org/>).

**Absence of Heterozygosity (AOH) Calculation.** To calculate genomic intervals and total genomic content of absence of heterozygosity (AOH) in each consanguineous individual, and as a surrogate measure for Runs-of-Homozygosity (ROH) Identity-by-descent (IBD), we used BafCalculator on unphased exome data^9^. For all SNVs that passed quality filters in a single variant call file, we extracted B-allele frequency (i.e. ratio of variant reads/total reads) and transformed this ratio by subtracting 0.5 and taking the absolute value for each data point. Values greater than 0.47 were regarded as homozygous variants corresponding to either alternative or reference alleles, whereas lower values were regarded as heterozygous variants. Transformed B-allele frequency data were then processed by circular binary segmentation implemented in the DNAcopy R Bioconductor package.

**Reverse transcriptase PCR (RT-PCR).** Blood was collected from the carrier mother of Individual 16 using PAXgene blood RNA tubes. Whole blood RNA was extracted using PAXgene blood RNA kit. RT-PCR was performed using primers targeting wild-type FLVCR1 cDNA, and PCR products were analysed by agarose gel electrophoresis. PCR products were sequenced by Sanger sequencing.

**Splicing Assay.** A minigene splicing assay was performed using a mini-gene split GFP construct^10^, in which N and C-terminal parts of the *GFP* gene were separated by *SMN1* introns 7 and 8 (NM_000344). Reference and mutated gene fragments (400-600bp) were synthetised (TWIST Bioscience, USA) and cloned into the mini-gene construct by Gibson Assembly (New England Biolabs). After Sanger sequencing verification of all constructs they were transfected into the HEK293 cells (Lipofectamine 3000, Thermo Fisher Scientific). Forty-eight hours post transfection, the cells were collected and total RNA was extracted (RNAeasy Mini Kit, Qiagen). After reverse transcription using random hexamers (SuperScript IV Synthesis Kit, Thermo Fisher Scientific), the minigene transcripts were amplified from the cDNA using primers specific to the split GFP fragments (F: 5’-CACACTGGTGACAACATTTACATAC-3’; R: 5’- GAAATCGTGCTGTTTCATGTGATC-3’). The PCR products were revealed on a 2% agarose gel and Sanger sequenced. If multiple bands were present they were gel extracted (Zymoclean Gel DNA Recovery Kit, Zymo Research) and Sanger sequenced. Alternatively column purified PCR products (DNA Clean & Concentrator-5, Zymo Research) were amplicon sequenced with next generation sequencing (Azenta Life Sciences). The gene sequence fragments used in this assay are available in **Supplemental Table 3**.

**Cell culture.** Human embryonic kidney (HEK293) cells were maintained in Dulbecco’s Modified Eagle Medium (DMEM) (Thermo Fisher Scientific) supplemented with 10% fetal bovine serum (Thermo Fisher Scientific) and 1% Penicillin-Streptomycin (Thermo Fisher Scientific). Cells were cultivated in an incubator at 37°C with 5% CO2.

**Plasmids and mutagenesis.** Human *FLVCR1* (*hFLVCR1*) plasmid was described previously. cDNA of long isoform of *hFLVCR1* (NM_014053.4) was cloned into pcDNA3.1 for overexpression. To generate missense mutations, *hFLVCR1* cDNA was used a template for site-directed mutagenesis via Polymerase Chain Reaction. The cDNA containing each missense mutation was cloned into pcDNA3.1 and confirmed by Sanger sequencing.

**Antibodies.** In-house polyclonal antibodies against human FLVCR1 were raised in rabbits against the 13 amino acids KMVMLSKQSESAI at the C-terminus. Other antibodies used were commercially available.

**Immunoblotting.** HEK293 cells were lysed with RIPA buffer (25 mM Tris pH 7–8, 150 mM NaCl, 0.1% SDS, 0.5% sodium deoxycholate 0.5% Triton X-100) containing protease inhibitor (Roche, 11836170001) and centrifuged at 16,000 g at 4°C to obtain protein lysates. Protein concentration was quantified via Pierce BCA protein assay (Thermo Fisher Scientific) following manufacturer’s protocol. 20 μg of protein was resolved on a 10% SDS–polyacrylamide gel electrophoresis (PAGE) gel and transferred onto a 0.45 μm nitrocellulose membrane. Membranes were blocked for in 5% milk in tris-buffered saline with tween-20 for 1 hour and incubated overnight with rabbit anti-human FLVCR1 primary antibody (1:500) or Mouse anti-GAPDH primary antibody (1:40000) (Santa Cruz, sc-32233) at 4°C, followed by incubation with Goat anti-rabbit IRDye® 800CW (1:10000) (LI-COR Biosciences, 926-32213) or IRDye® 680LT Goat anti-Mouse IgG (H+L) (1:10000) (LI-COR Biosciences, 926-68020) secondary antibody for 1 hour at room temperature. PageRuler Plus Prestained Protein Ladder (Thermo Scientific, 26620) was used as protein size standard. Membranes were imaged by ChemiDoc MP Imaging system (Bio-Rad).

**Immunofluorescence staining.** HEK293 cells were co-transfected using *hFLVCR1* or mutant plasmids with the plasma membrane marker pCAG-mGFP plasmid using Lipofectamine 2000 (Thermo Fisher Scientific). 24 hours post-transfection, cells fixed with 4% paraformaldehyde for 15 minutes, permeabilized with 0.1% Triton-X in PBS for 10 minutes and blocked with 5% Natural Goat Serum (NGS) in 0.1% Triton-X for 15 minutes. The cells were incubated with rabbit anti-human FLVCR1 primary antibody (1:200) (in 5% Natural Goat Serum (NGS) in 0.1% Triton-X in PBS) for 1 hour, washed with PBS, followed by incubation with goat anti-rabbit IgG (H+L) Alexa Fluor® 555 conjugate secondary antibody (1:1000) (Abcam) for 1 hour. Cells were counterstained with Hoechst 33342 for 55 minutes to visualize the nuclei. Coverslips were mounted on a glass slide with mounting media (70% glycerol, 20mM Tris HCL, pH 6) and visualized using the LSM710 microscope (Carl Zeiss MicroImaging) with a 63x oil immersion objective. Immunofluorescence images were analysed using the LSM Image Browser software (Carl Zeiss MicroImaging) and processed via Adobe Photoshop CS6.

**Transport assays.** HEK293 cells were co-transfected using *hFLVCR1* or mutant plasmids with human Choline kinase alpha (*hCHKA*) plasmid. Mock was transfected with *hCHKA* plasmid only and used as control. 20-30 hours post-transfection, HEK293 cells were incubated with 100µM [^3^H] choline in DMEM containing 10% FBS for 1 hour at 37°C. The cells were washed twice with cold plain DMEM. Cell pellets were lysed in RIPA buffer and transferred to scintillation vials for quantification of radioactive signal using Tricarb liquid scintillation counter. Transport activity of hFLVCR1 mutants was expressed percentage of wild-type hFLVCR1 protein. Each symbol on graph represents one replicate.

**Multiple sequence alignment.** The predicted amino acid sequence of Homo sapiens and Mus musculus FLVCR1 and FLVCR2 was extracted from the NCBI database with the following identifiers: Homo sapiens FLVCR1 (NP_054772.1); Mus musculus FLVCR1 (NP_001074728.1); Homo sapiens FLVCR2 (NP_060261.2); Mus musculus FLVCR2 (NP_663422.1). Multiple sequence alignment was done using ClustalW^11^.

**Statistical Analyses.** Data were analysed using GraphPad Prism 8. Statistical significance was determined using unpaired t test with Welch’s correction or one-way ANOVA. P-value < 0.05 was considered statistically significant.

**Supplemental Data and Figures:**

**Per medrxiv policies, autopsy reports for Individuals 11 and 17 are omitted but are available upon request to the corresponding authors**

**Supplemental Figure 1: Absence of heterozygosity (AOH) data from Individual 1 (I1), Family 1 is suggestive of a South Asian founder allele.**

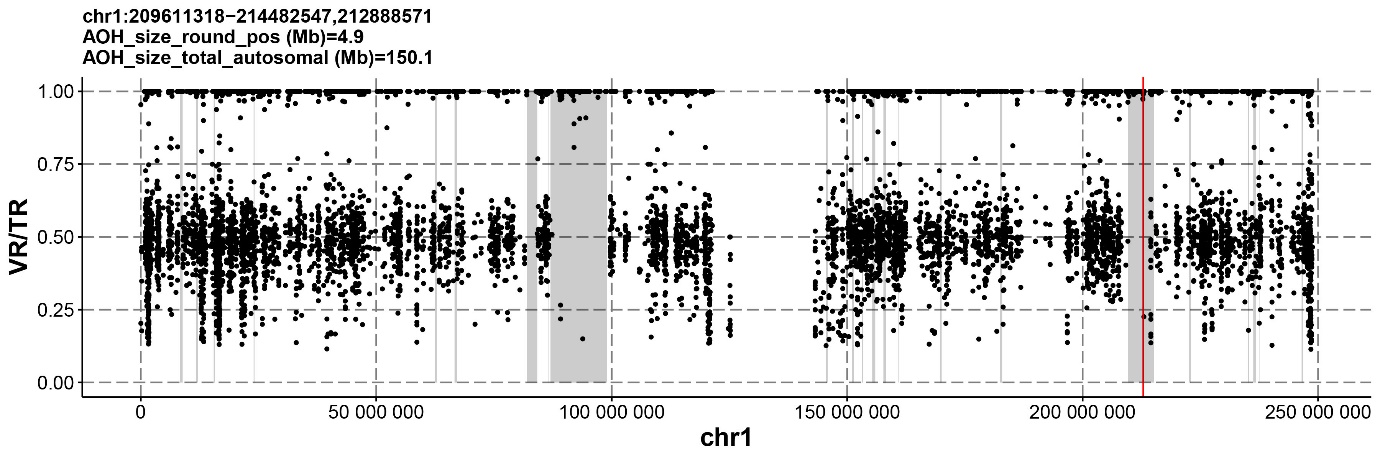

**Supplemental Fig 1:**

B-allele frequency for Individual 1 (I1) was calculated from exome variant data. Individual 1 is a male child of South Asian descent born to unrelated parents. A 4.9 Mb block for AOH on chromosome 1 (grey) was detected around the *FLVCR1* variant c.1390G>A p.G494S. Total autosomal AOH was calculated at 150.1 Mb. The homozygous *FLVCR1* variant c.1390G>A was also identified in Individual 8, an unrelated child of South Asian descent from a non-consanguineous family. These observations combined with the presence of an AOH block surrounding the *FLVCR1* variant in individual 1 is suggestive of a South Asian founder allele.

**Supplemental Figure 2: Photographs and brain magnetic resonance imaging findings in Family 17**

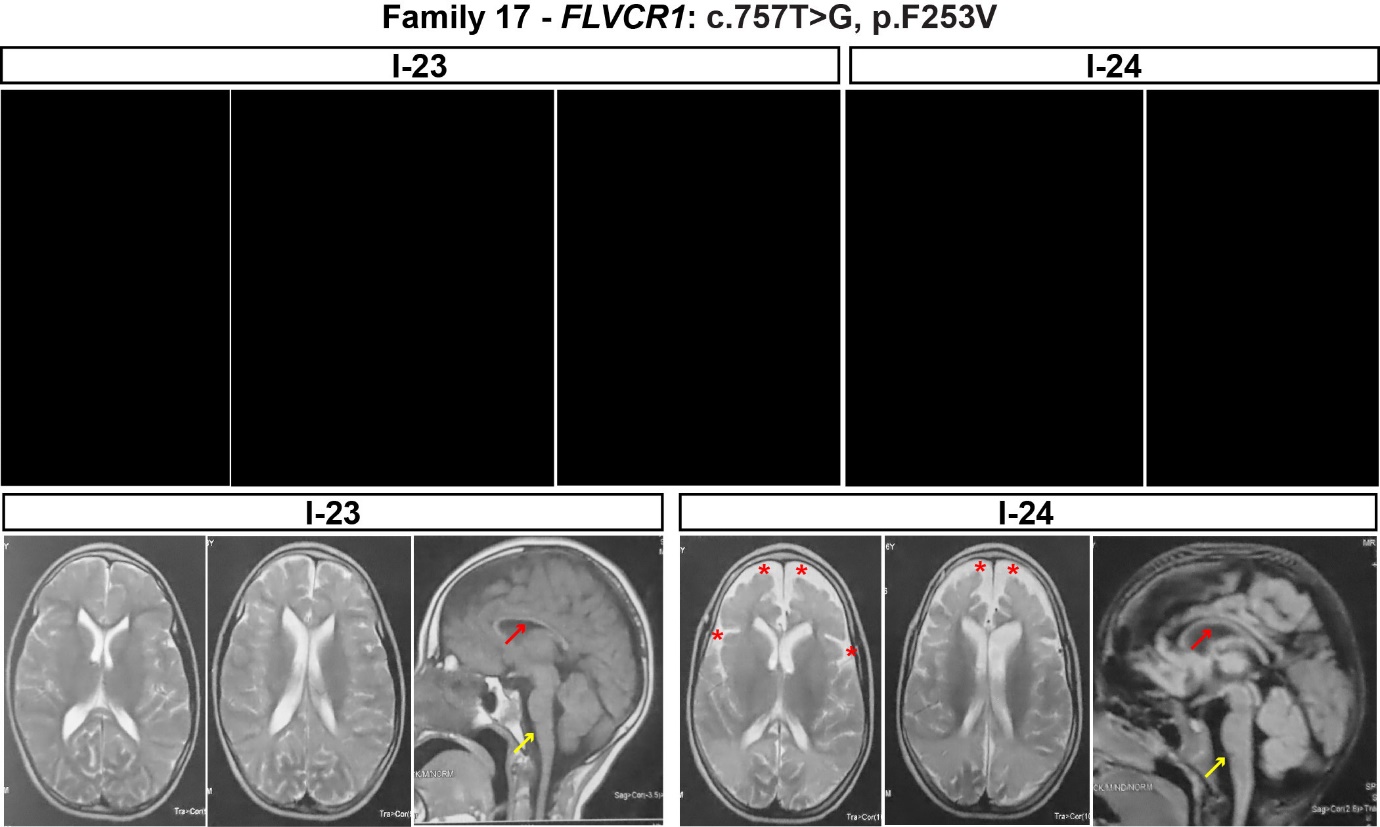

**Supplemental Fig S2:**

The younger sibling in Family 17 (I-24) exhibited more severe developmental impairment than his older brother (I-23). While I-23 had delayed acquisition of independent walking, I-24 remains non-ambulatory but can stand with support and sit independently. Both are non-verbal. Brain MRI of I-24 showed a greater degree of increased extra-fluid space enlargement and white matter reduction than I-23 (bottom row)

**Top row** - Photographs of Individual 23 demonstrate microcephaly and dysmorphic features including long face, triangular broad nose, prominent columella, short philtrum, macrostomia, everted lower lip, prominent teeth, and low-set ears. Photographs of Individual 24 show microcephaly and long face, bitemporal hollowing, prominent nasal columella, short philtrum, large mouth, prominent mandible and low-set ears. **Per medrxiv requirements, patient photographs have been removed but are available upon request to the corresponding authors.**

**Bottom row** – Magnetic resonance imaging of Individual 23 and 24 shows prominent ventricles, corpus callosum thinning (red arrow), increased extra-axial fluid spaces (red asterisks), reduced white matter volume, and brainstem and pontine thinning (yellow arrow).

**Supplemental Figure 3: Multiple sequence alignment of human and mouse *FLVCR1* and *FLVCR2* in and summary of *FLVCR1* missense variants.**
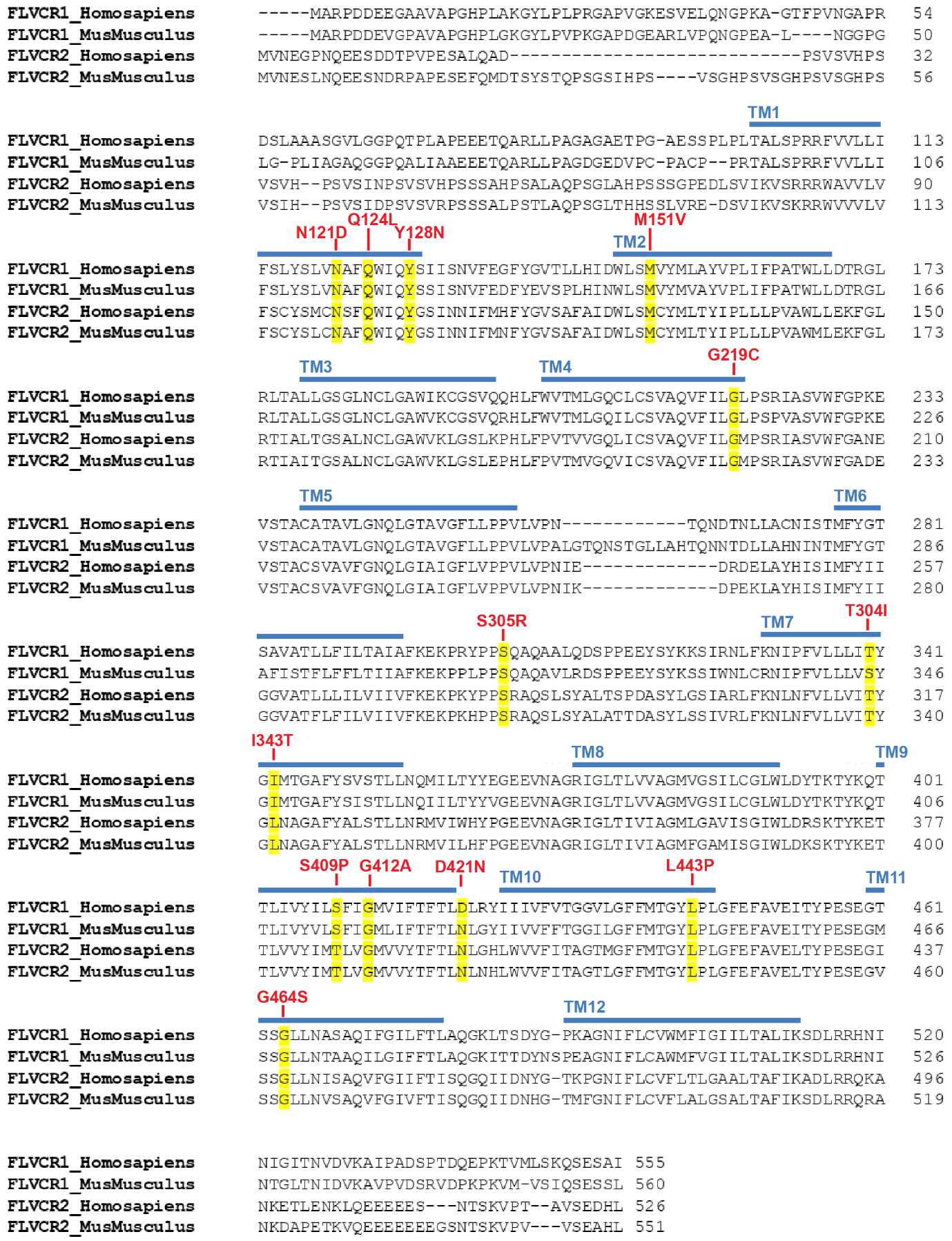

**Supplemental Fig 3:**

FLVCR1 is a conserved protein from flies which has 12 predicted transmembrane (TM) domains (TM1-12). Blue region indicates the transmembrane domains. Amino acid sequences of *Homo sapiens* and *Mus musculus* FLVCR1 and FLVCR2 were aligned with ClustalW. Numbers on the left indicate amino acid positions, dashes indicate gaps. Missense variants of FLVCR1 observed in patients in this study are highlighted in yellow and the specific amino acid change are indicated in red letters. Most missense variants are conserved across the species.

**Supplemental Figure 4: Splicing studies of *FLVCR1*: c.884-3C>G**

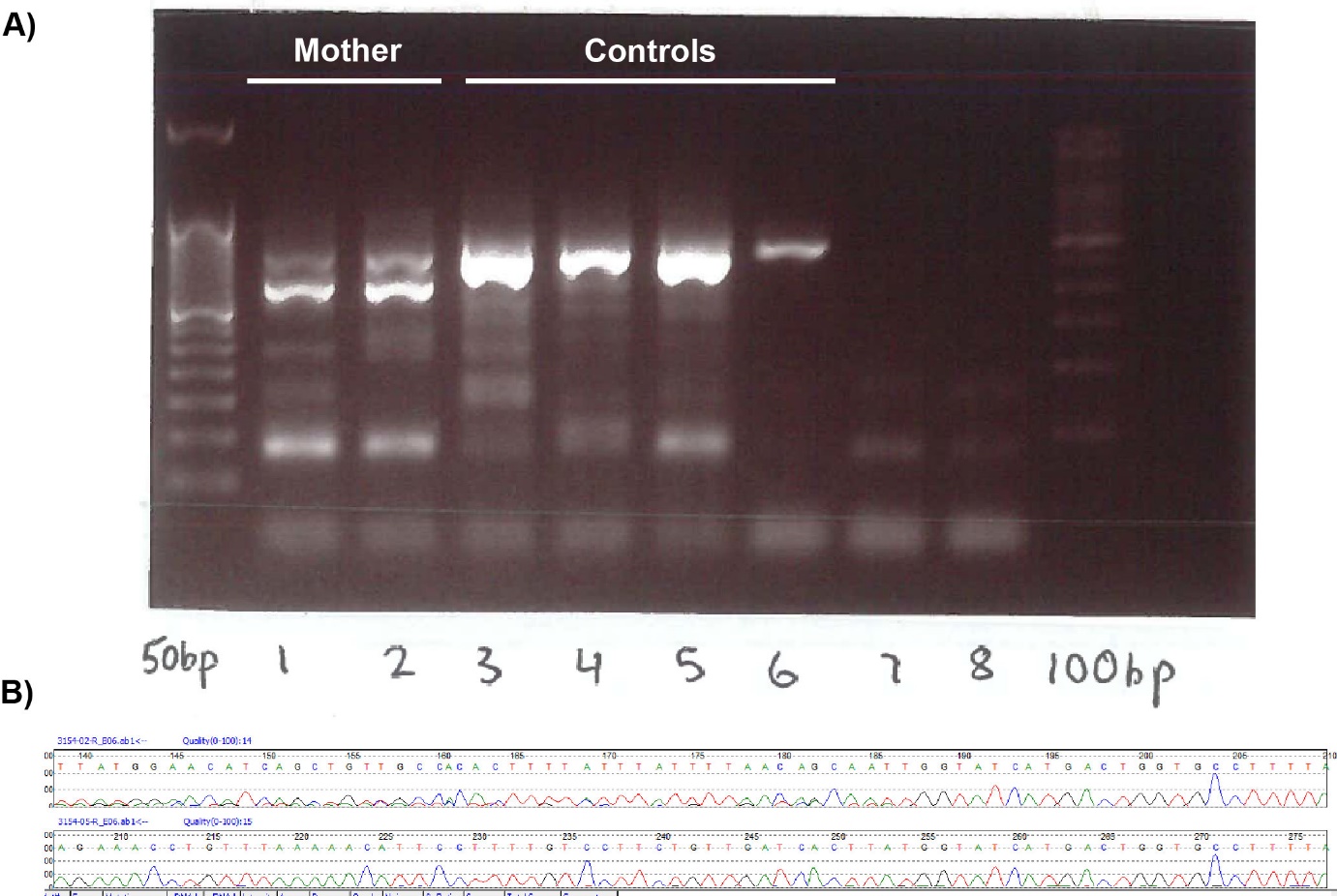

**Supplemental Fig 4:**

**A)** Agarose gel showing RT-PCR results from whole blood RNA from mother of individual 16 who carries the *FLVCR1* c.884-3C>G variant (lanes 1,2), controls (lanes 3-6), and negative controls (lanes 7,8).

**B)** Sanger sequencing performed on RT-PCR bands from Fig. S2A. Top row shows Sanger sequencing results from mother’s sample, whereas bottom row shows Sanger sequencing results from control sample. The two bands visible in the mother’s samples give the appearance of a frameshift on Sanger sequencing while only a single tracing corresponding to wild-type FLVCR1 is visible in the control sample. Analysis of the mother’s Sanger sequencing demonstrated a heterozygous inframe deletion of exon 3: r.884_1024del p.A295_Y341del.

**Supplemental Figure 5**: Mini-gene splicing assay demonstrates splicing defects secondary to *FLVCR1* single nucleotide variants and indels

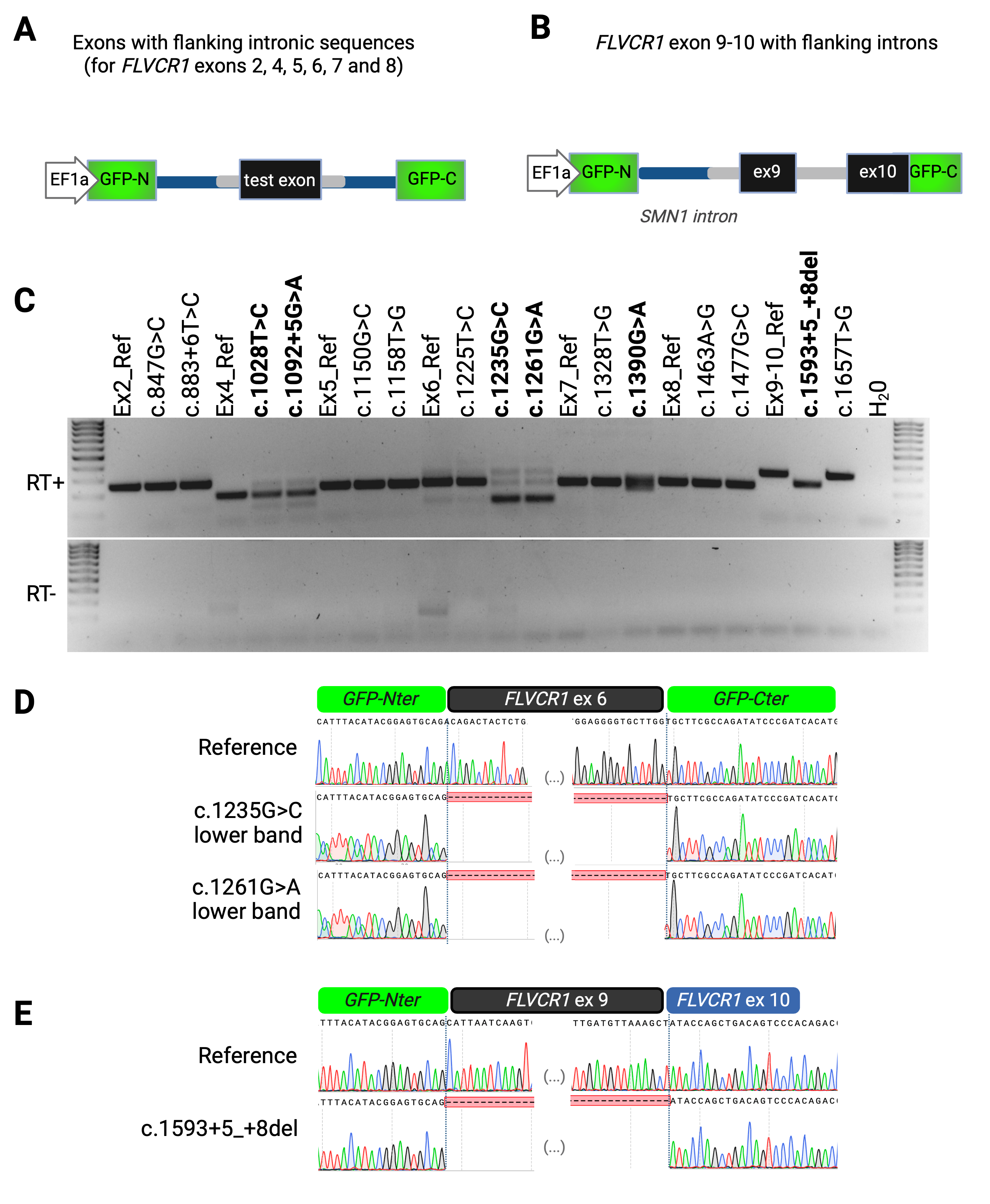

**Supplemental Fig 5:**

**A)** Mini-gene splicing assay strategy for *FLVCR1* variants within exons 2, 4, 5, 6, 7, or 8. The exon of interest including flanking *FLVCR1* intronic sequences (shown in gray) were cloned into the mini-gene construct within *SMN1* introns 7 and 8 (shown in blue). Exons are shown in black and GFP is shown in green.

**B)** Mini-gene splicing assay strategy for *FLVCR1* variants within exons 9 or 10. Exons 9 and 10 were cloned into the mini-gene construct as shown. Flanking *FLVCR1* intronic sequences are shown in gray, *SMN1* introns 7 and 8 are shown in blue, exons are shown in black, and GFP is shown in green.

**C)** Mini-gene splicing assay strategy for *FLVCR1* variants within exons 9 or 10. Exons 9 and 10 were cloned into the mini-gene construct as shown. Flanking *FLVCR1* intronic sequences are shown in gray, *SMN1* introns 7 and 8 are shown in blue, exons are shown in black, and GFP is shown in green.

**D)** Results of RT-PCR performed on either reference sequences (labelled Ex2_Ref, Ex4_Ref, etc) or variant sequences (top gel). Variants which impact splicing are shown in bold. Reverse transcriptase negative control is shown in the bottom gel.

**E)** Sanger sequencing results of the reference exon 6 as well as dominant, lower molecular weight RT-PCR bands for *FLVCR1*:c.1235G>C and c.1261G>A. Both *FLVCR1*:c.1235G>C and c.1261G>A demonstrate exon 6 skipping.

**F)** Sanger sequencing results of reference exons 9 and 10 as well as the lower molecular weight RT-PCR band for *FLVCR1*:c.1593+5_+8del. c.1593+5_+8del causes exon 9 skipping.

**Supplemental Figure 6**: *FLVCR1* exon 4 variants c.1028T>C and c.1092+5G>A induce exon skipping
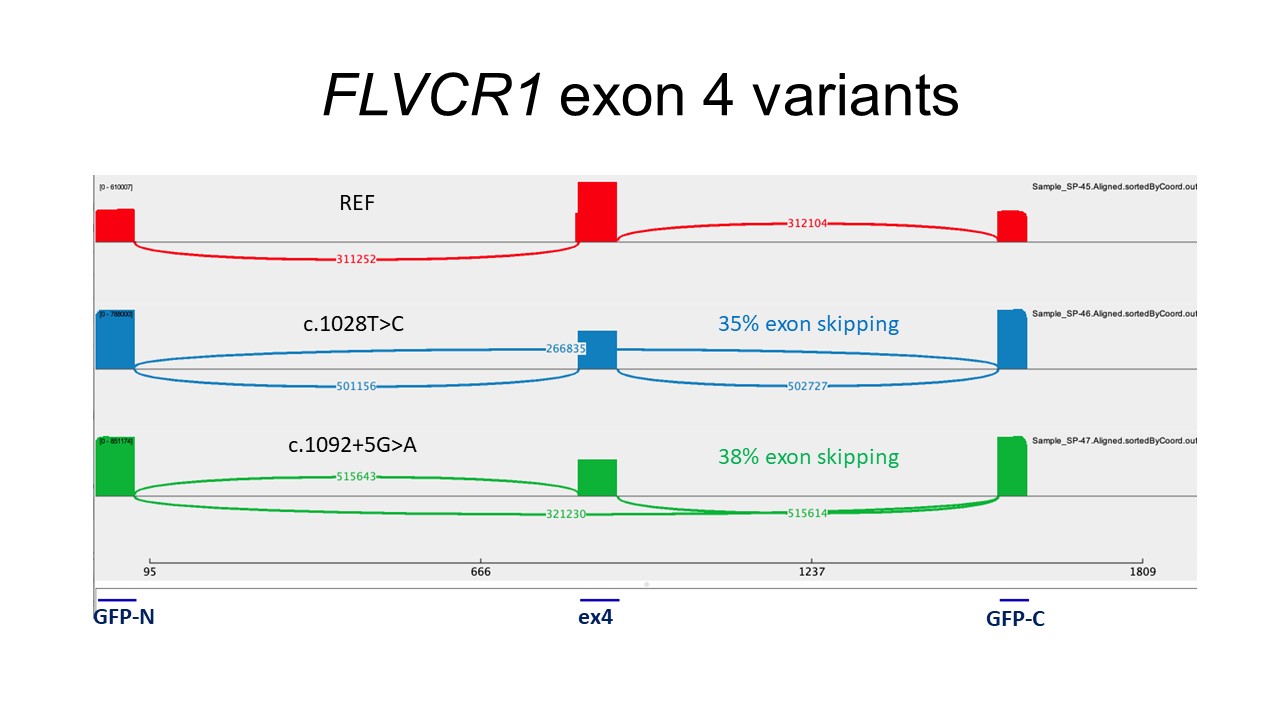

**Supplemental Fig 6:**

Sashimi plots generated from next generation sequencing of RNA from HEK293 cells transfected with constructs containing the reference sequence of *FLVCR1* exon 4 and flanking intronic regions (Ex4_Ref, red), *FLVCR1*: c.1028T>C (blue), or *FLVCR1*: c.1092+5G>A (green).

**Supplemental Figure 7**: *FLVCR1* exon 6 variants c.1235G>C and c.1261G>A induce exon skipping
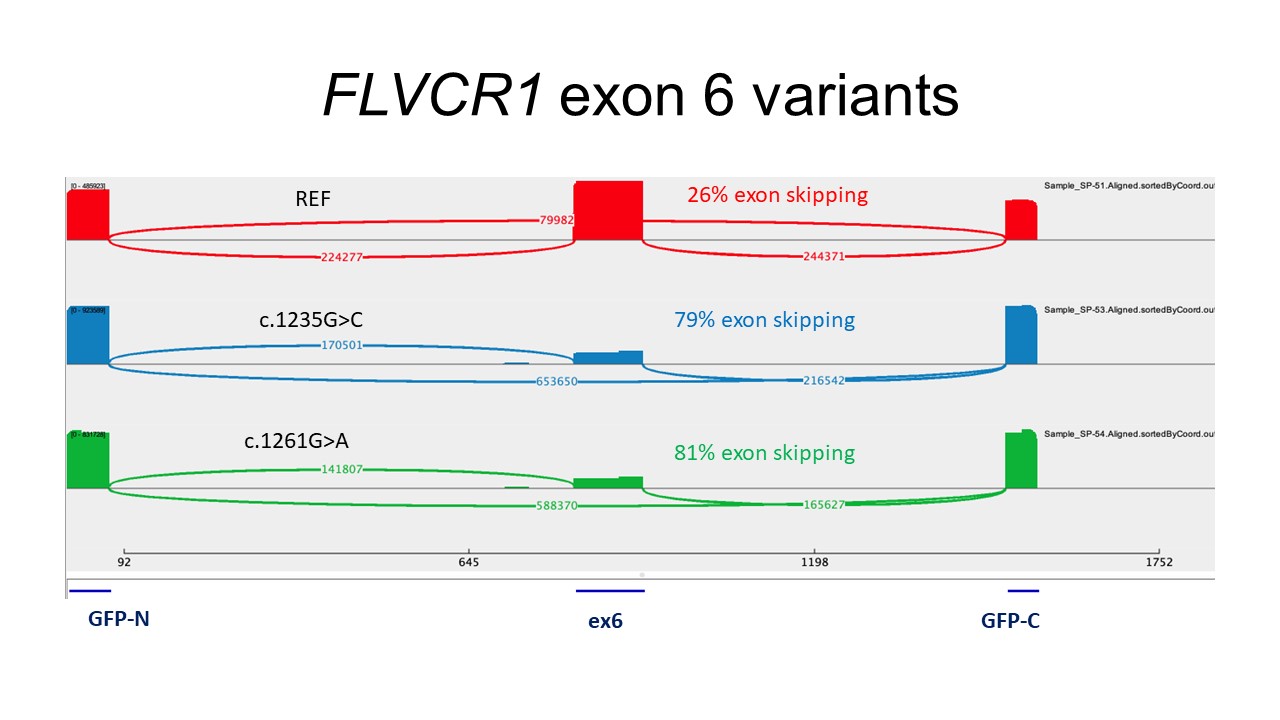

**Supplemental Fig 7:**

Sashimi plots generated from next generation sequencing of RNA from HEK293 cells transfected with constructs containing the reference sequence of *FLVCR1* exon 6 and flanking intronic regions (Ex6_Ref, red), *FLVCR1*: c.1235G>C (blue), or *FLVCR1*: c.1261G>A (green).

**Supplemental Figure 8**: *FLVCR1* exon 7 variants c.1390G>A increase the utilization of an alternative donor site
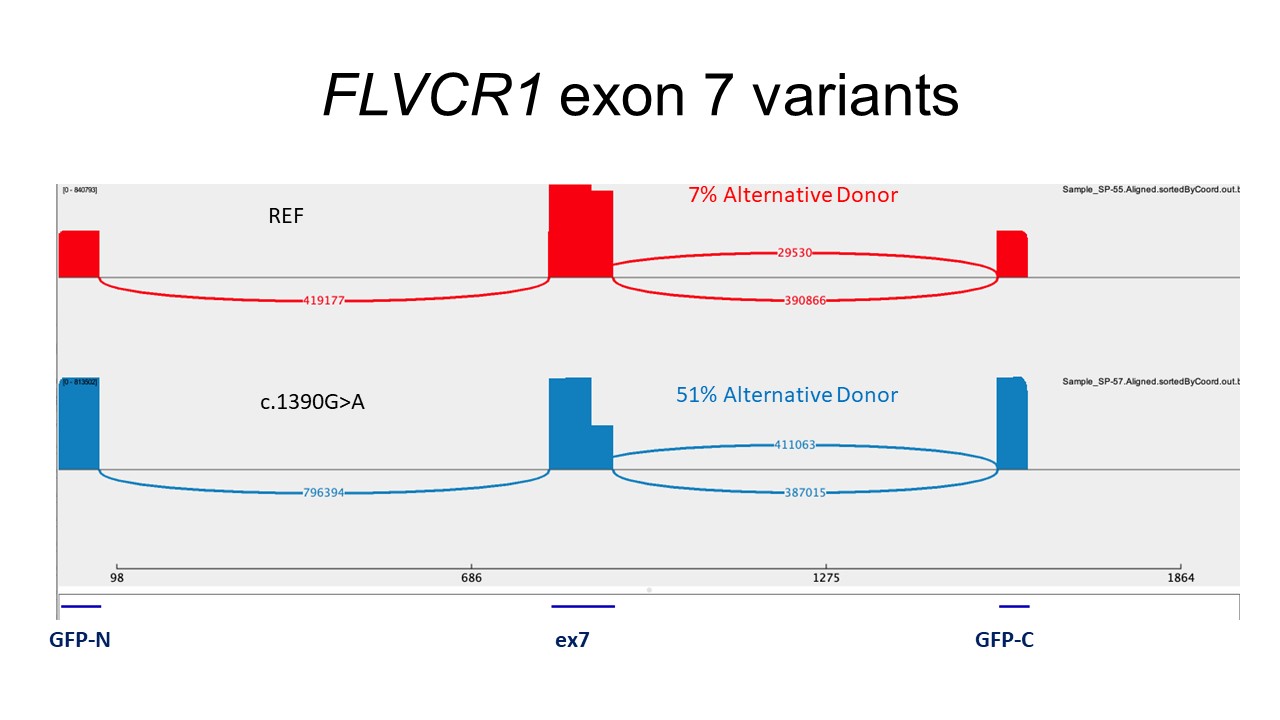

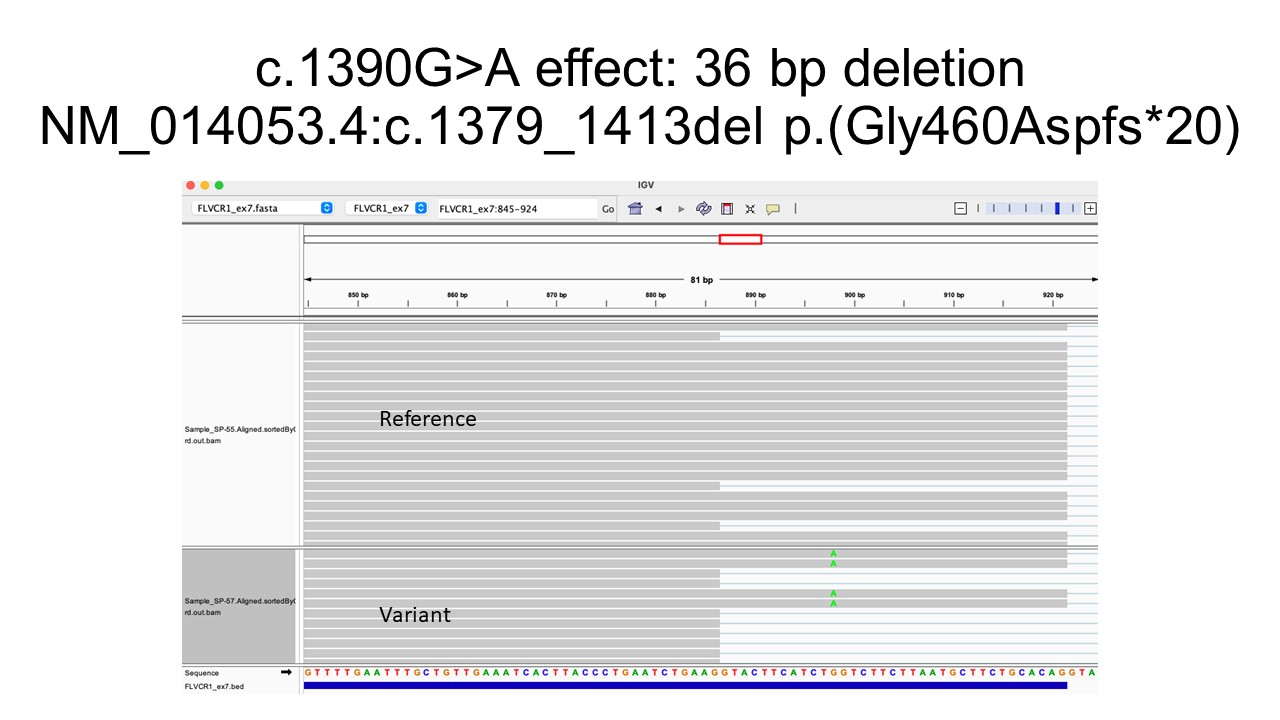

**Supplemental Fig 8:**

**Top figure:** Sashimi plots generated from next generation sequencing of RNA from HEK293 cells transfected with constructs containing the reference sequence of *FLVCR1* exon 7 and flanking intronic regions (Ex7_Ref, red) or *FLVCR1*: c.1390G>A (blue).

**Bottom figure:** Visualization of next generation sequencing data in Integrative Genomics Viewer demonstrating a 36 bp deletion resulting from the alternative donor site.

**Supplemental Figure 9**: *FLVCR1* exon 9 variant c.1593+5_+8del induce exon skipping
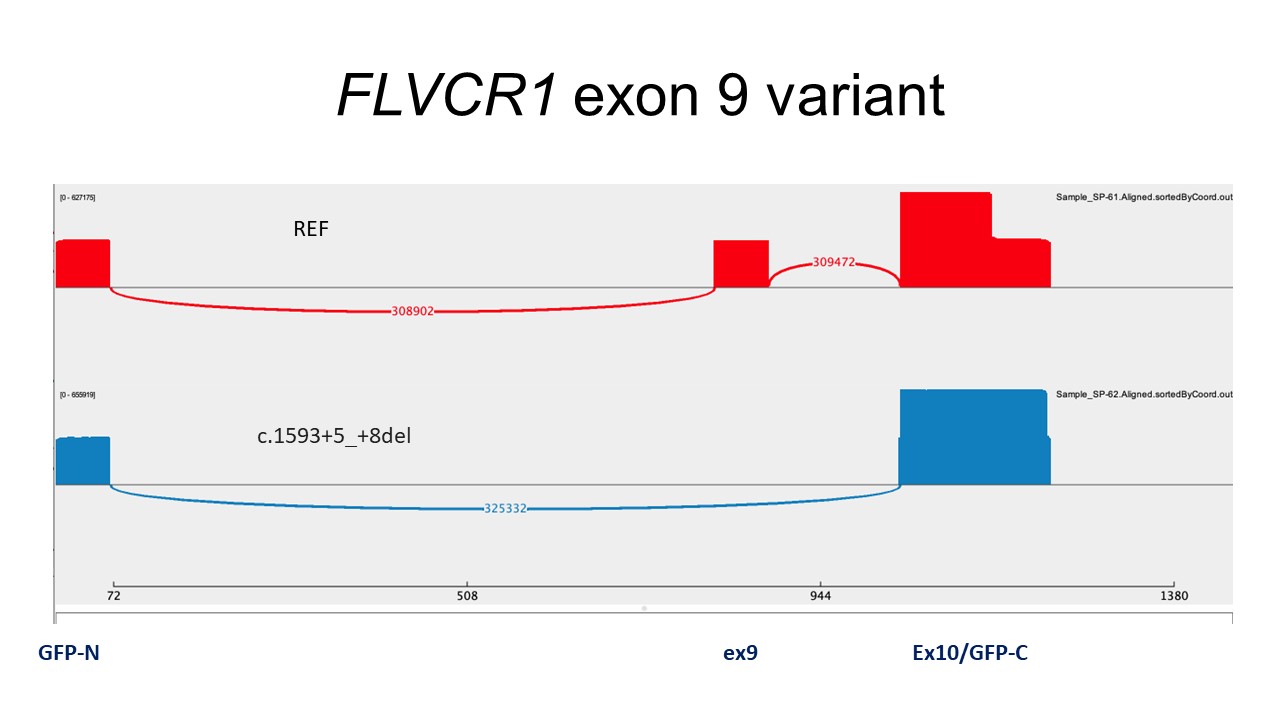

**Supplemental Fig 9:**

Sashimi plots generated from next generation sequencing of RNA from HEK293 cells transfected with constructs containing the reference sequence of *FLVCR1* exon 9 and flanking intronic regions (Ex6_Ref, red) or *FLVCR1*: c.1593+5_+8del (blue).

**Supplemental Table 1: Summary of all previously reported and novel *FLVCR1* variants and associated phenotypes (attached .xls file)**

**Supplemental Table 2: Choline and ethanolamine transport activity for each FLVCR1 missense variant examined in this study**

| Supplemental Table 2: Summary of choline and ethanolamine import activity of FLVCR1 missense variants relative to WT (%) in patients from this study and the literature. | | | | | | |
| --- | --- | --- | --- | --- | --- | --- |
| Choline import activity | | | | | | |
| Individual(s) | **Reported symptoms** | **Allele 1** | **Allele 2** | **Activity Allele 1** | **Activity Allele 2** | **References** |
| F2, I-2 | Severe NDD | L443P | - | 29.52% | 29.52% | This study |
| F6, I-6,I-7 | Severe NDD | G412A | - | 0% | 0% | This study |
| MPV case |  | M151V | - | 93.77% | 93.77% | This study |
| F14, I-20; F17 I-23 | HSAN, RP | I343T | - | 41.18% | 41.18% | This study |
| F1, I-1 | Severe NDD | G464S | - | 17.76% | 17.76% | This study |
| F5, I-5 | Severe NDD | S305R | - | 55.38% | 55.38% | This study |
| F15, I-21; F16, I-22 | PCARP, HSP | Y128N | - | 44.61% | 44.61% | This study |
| F3, I-3 | Severe NDD | D421N | L390* | 99.74% | - | This study |
| F8, I-9, I-10, I-11 | Severe NDD | T340I | S409P | 8.87% | 39.23% | This study |
|  | HSAN, ID | Q124L | G219C | 17.79% | 26.82% | ^12^ |
| Ethanolamine import activity | | | | | | |
| F2, I-2 | Severe NDD | L443P | - | 6.02% | 6.02% | This study |
| F6, I-6,I-7 | Severe NDD | G412A | - | 0% | 0% | This study |
| MPV case |  | M151V | - | 91.30% | 91.30% | This study |
| F14, I-20; F17 I-23 | HSAN, RP | I343T | - | 48.80% | 48.80% | This study |
| F1, I-1 | Severe NDD | G464S | - | 38.80% | 38.80% | This study |
| F5, I-5 | Severe NDD | S305R | - | 8.46% | 8.46% | This study |
| F15, I-21; F16, I-22 | PCARP, HSP | Y128N | - | 29.64% | 29.64% | This study |
| F3, I-3 | Severe NDD | D421N | L390* | 97.77% | - | This study |
| F8, I-9, I-10, I-11 | Severe NDD | T340I | S409P | 11.30% | 2.70% | This study |
|  | HSAN, ID | Q124L | G219C | 6.62% | 36.77% | ^12^ |

**Supplemental Table 3: Gene fragment sequences used in mini-gene splicing assay (attached .csv file)**

**Supplemental References:**

1.	Sobreira, N., Schiettecatte, F., Valle, D., and Hamosh, A. (2015). GeneMatcher: a matching tool for connecting investigators with an interest in the same gene. Hum Mutat *36*, 928–930. 10.1002/humu.22844.

2.	Wohler, E., Martin, R., Griffith, S., Rodrigues, E. da S., Antonescu, C., Posey, J.E., Coban-Akdemir, Z., Jhangiani, S.N., Doheny, K.F., Lupski, J.R., et al. (2021). PhenoDB, GeneMatcher and VariantMatcher, tools for analysis and sharing of sequence data. Orphanet Journal of Rare Diseases *16*, 365. 10.1186/s13023-021-01916-z.

3.	Calame, D.G., Bakhtiari, S., Logan, R., Coban-Akdemir, Z., Du, H., Mitani, T., Fatih, J.M., Hunter, J.V., Herman, I., Pehlivan, D., et al. (2021). Biallelic loss-of-function variants in the splicing regulator NSRP1 cause a severe neurodevelopmental disorder with spastic cerebral palsy and epilepsy. Genet Med *23*, 2455–2460. 10.1038/s41436-021-01291-x.

4.	Calame, D.G., Guo, T., Wang, C., Garrett, L., Jolly, A., Dawood, M., Kurolap, A., Henig, N.Z., Fatih, J.M., Herman, I., et al. (2023). Monoallelic variation in DHX9, the gene encoding the DExH-box helicase DHX9, underlies neurodevelopment disorders and Charcot-Marie-Tooth disease. The American Journal of Human Genetics *110*, 1394–1413. 10.1016/j.ajhg.2023.06.013.

5.	Bainbridge, M.N., Wang, M., Wu, Y., Newsham, I., Muzny, D.M., Jefferies, J.L., Albert, T.J., Burgess, D.L., and Gibbs, R.A. (2011). Targeted enrichment beyond the consensus coding DNA sequence exome reveals exons with higher variant densities. Genome Biol *12*, R68. 10.1186/gb-2011-12-7-r68.

6.	Challis, D., Yu, J., Evani, U.S., Jackson, A.R., Paithankar, S., Coarfa, C., Milosavljevic, A., Gibbs, R.A., and Yu, F. (2012). An integrative variant analysis suite for whole exome next-generation sequencing data. BMC Bioinformatics *13*, 8. 10.1186/1471-2105-13-8.

7.	Reid, J.G., Carroll, A., Veeraraghavan, N., Dahdouli, M., Sundquist, A., English, A., Bainbridge, M., White, S., Salerno, W., Buhay, C., et al. (2014). Launching genomics into the cloud: deployment of Mercury, a next generation sequence analysis pipeline. BMC Bioinformatics *15*, 30. 10.1186/1471-2105-15-30.

8.	Eldomery, M.K., Coban-Akdemir, Z., Harel, T., Rosenfeld, J.A., Gambin, T., Stray-Pedersen, A., Küry, S., Mercier, S., Lessel, D., Denecke, J., et al. (2017). Lessons learned from additional research analyses of unsolved clinical exome cases. Genome Medicine *9*, 26. 10.1186/s13073-017-0412-6.

9.	Karaca, E., Posey, J.E., Akdemir, Z.C., Pehlivan, D., Harel, T., Jhangiani, S.N., Bayram, Y., Song, X., Bahrambeigi, V., Yuregir, O.O., et al. (2018). Phenotypic expansion illuminates multilocus pathogenic variation. Genetics in Medicine *20*, 1528–1537. 10.1038/gim.2018.33.

10.	Scott, H.A., Place, E.M., Harper, E., Mehrotra, S., Cmg, B., Huckfeldt, R., Comander, J., Pierce, E.A., and Bujakowska, K.M. (2023). A high throughput splicing assay to investigate the effect of variants of unknown significance on exon inclusion. Preprint at medRxiv, 10.1101/2022.11.30.22282952 10.1101/2022.11.30.22282952.

11.	Sievers, F., Wilm, A., Dineen, D., Gibson, T.J., Karplus, K., Li, W., Lopez, R., McWilliam, H., Remmert, M., Söding, J., et al. (2011). Fast, scalable generation of high-quality protein multiple sequence alignments using Clustal Omega. Molecular Systems Biology *7*, 539. 10.1038/msb.2011.75.

12.	Suthar, R., Sharawat, I.K., Eggermann, K., Padmanabha, H., Saini, A.G., Bharti, B., Kurth, I., Singhi, P., and Sankhyan, N. (2022). Hereditary Sensory and Autonomic Neuropathy: A Case Series of Six Children. Neurology India *70*, 231. 10.4103/0028-3886.338691.
